# Association between COVID-19 mRNA vaccination and COVID-19 illness and severity during Omicron BA.4 and BA.5 sublineage periods

**DOI:** 10.1101/2022.10.04.22280459

**Authors:** Ruth Link-Gelles, Matthew E. Levy, Karthik Natarajan, Sarah E. Reese, Allison L. Naleway, Shaun J. Grannis, Nicola P. Klein, Malini B. DeSilva, Toan C. Ong, Manjusha Gaglani, Emily Hartmann, Monica Dickerson, Edward Stenehjem, Anupam B. Kharbanda, Jungmi Han, Talia L. Spark, Stephanie A. Irving, Brian E. Dixon, Ousseny Zerbo, Charlene E. McEvoy, Suchitra Rao, Chandni Raiyani, Chantel Sloan-Aagard, Palak Patel, Kristin Dascomb, Anne-Catrin Uhlemann, Margaret M. Dunne, William F. Fadel, Ned Lewis, Michelle A. Barron, Kempapura Murthy, Juan Nanez, Eric P. Griggs, Nancy Grisel, Medini K. Annavajhala, Akintunde Akinseye, Nimish R. Valvi, Kristin Goddard, Mufaddal Mamawala, Julie Arndorfer, Duck-Hye Yang, Peter J. Embí, Bruce Fireman, Sarah W. Ball, Mark W. Tenforde

**Author notes:** **Corresponding Author:** Ruth Link-Gelles, PhD, MPH. US Centers for Disease Control and Prevention. Mail: 1600 Clifton Rd, Mailstop H24-5, Atlanta, Georgia 30329. **Disclaimer:** The findings and conclusions in this report are those of the authors and do not necessarily represent the official position of the Centers for Disease Control and Prevention.

## Abstract

**Importance:** Recent sublineages of the SARS-CoV-2 Omicron variant, including BA.4 and BA.5, may be associated with greater immune evasion and less protection against COVID-19 following vaccination.

**Objective:** To evaluate the association between COVID-19 mRNA vaccination with 2, 3, or 4 doses among immunocompetent adults and the risk of medically attended COVID-19 illness during a period of BA.4/BA.5 predominant circulation; to evaluate the relative severity of COVID-19 in hospitalized cases across Omicron BA.1, BA.2/BA.2.12.1, and BA.4/BA.5 sublineage periods.

**Setting, Design and Participants:** Test-negative study of adults with COVID-19-like illness (CLI) and molecular testing for SARS-CoV-2 conducted in 10 states from December 16, 2021, to August 20, 2022.

**Exposure:** mRNA COVID-19 vaccination.

**Main Outcomes and Measures:** Emergency department/urgent care encounters, hospitalizations, and admission to the intensive care unit (ICU) or in-hospital death. The adjusted odds ratio (OR) for the association between prior vaccination and medically attended COVID-19 was used to estimate VE, stratified by care setting and vaccine doses (2, 3, or 4 doses vs 0 doses as reference group). Among hospitalized case-patients, demographic and clinical characteristics and in-hospital outcomes including ICU admission and death were compared across sublineage periods.

**Results:** Between June 19 – August 20, 2022, 82,229 ED/UC and 21,007 hospital encounters were included for the BA.4/BA.5 vaccine effectiveness analysis. Among adults hospitalized with CLI, the adjusted odds ratio (OR) was 0.75 (95% CI: 0.68-0.83) for receipt of 2 vaccine doses at ≥150 days after receipt, 0.32 (95% CI: 0.20-0.50) for a third dose 7-119 days after receipt, and 0.64 (95% CI: 0.58-0.71) for a third dose ≥120 days (median 235 days) after receipt for cases vs controls. For COVID-19-associated hospitalization, among patients ages ≥65 years 7-59 and ≥60 days (median 88 days) after a fourth dose, ORs were 0.34 (95% CI: 0.25-0.47) and 0.43 (95% CI: 0.34-0.56), respectively. Among hospitalized cases, ICU admission and/or in-hospital death occurred in 21.4% during the BA.1 vs 14.7% during the BA.4/BA.5 period (standardized mean difference: 0.17).

**Conclusion:** VE against medically attended COVID-19 illness decreased over time since last dose; receipt of one or two booster doses increased effectiveness over a primary series alone.

**KEY POINTS:** 

**Question:** What is the association between receipt of first-generation COVID-19 mRNA vaccines and medically attended COVID-19 during Omicron BA.4/BA.5 sublineage predominance?

**Findings:** This test-negative analysis included 82,229 emergency department or urgent care encounters and 21,007 hospitalizations for COVID-19-like illness. Among hospitalized patients, the likelihood of recent vaccination (7-119 days) with 3 mRNA vaccine doses (vs unvaccinated) was significantly lower (odds ratio, 0.32) in cases than SARS-CoV-2-negative controls, but with lower associated protection ≥120 days post-vaccination (odds ratio, 0.64).

**Meaning:** First-generation COVID-19 vaccines were associated with protection against COVID-19 during the Omicron BA.4/BA.5 sublineage-predominant periods but this declined over time.

## INTRODUCTION

COVID-19 vaccines are estimated to have prevented tens of thousands of COVID-19-associated hospitalizations and deaths in the United States (US).^1^ Over the course of the pandemic, however, new SARS-CoV-2 variants have continued to emerge and evade vaccine-induced immunity.^2^ Following a Delta variant predominant period, the Omicron BA.1 sublineage became predominant in the United States by December 2021. Compared to earlier SARS-CoV-2 variants, BA.1 demonstrated increased transmissibility and immune evasion with a reduction in vaccine effectiveness (VE) offset by COVID-19 vaccine booster doses.^3,4^ Omicron has since diversified into additional sublineages, including several (eg, BA.2.12.1, BA.4, and BA.5) with greater immune escape potential compared to BA.1.^5,6^ BA.4 and BA.5 sublineages, which share a common spike protein, became the predominant circulating viruses in the US in June 2022.^7^

As new circulating variants emerge, ongoing monitoring of VE is critical for informing public health strategies and policies. COVID-19 VE estimation has become complex as additional vaccine booster doses are authorized, vaccine-induced protection wanes over time, new variants or subvariants have emerged, and a majority of the US population has experienced previous infection (57-94%, depending on source).^8-11,12^ In November 2021, all adults were recommended to receive a third (booster) vaccine dose after a two-dose primary series of mRNA vaccine; in March 2022, adults ≥50 years of age were recommended to receive a fourth dose (second booster) at least 4 months after dose 3.^13^ In September 2022, bivalent mRNA vaccine booster doses for all individuals ≥12 years of age (Pfizer-BioNTech) and adults ≥18 years of age (Moderna) were recommended at least 2 months after completing a primary series or receiving a booster dose.^14^ Like first-generation vaccines, bivalent vaccines contain an mRNA component targeting the ancestral virus in addition to a new component targeting theBA.4/BA.5 spike protein.

The objectives for this analysis were: 1) to estimate the association of first-generation mRNA vaccines (BNT162b2 from Pfizer-BioNTech and mRNA-1273 from Moderna) with medical encounters for COVID-19-associated illness during a period of BA.4/BA.5 Omicron sublineage predominance among adults without immunocompromising conditions and 2) to describe the epidemiology and severity of hospitalized COVID-19 cases during the Omicron BA.4/BA.5 predominant period compared to prior Omicron sublineage periods (BA.1 and BA.2/BA.2.12.1). Understanding changes in the epidemiology of COVID-19 and VE will inform interpretation of VE studies for recently authorized bivalent vaccines.

## METHODS

### Design and Setting

The VISION Network is a multistate collaboration between the U.S. Centers for Disease Control and Prevention (CDC) and health care systems with integrated medical, laboratory, and vaccination records. VISION performs serial assessments of COVID-19 VE in emergency department/urgent care (ED/UC) and hospital settings using the test-negative case-control design.^15^ Nine VISION sites in 10 states contributed data for this analysis (eTable 1), including 268 hospitals, 292 emergency departments, and 140 urgent care clinics.

For the VE analysis, events included ED/UC encounters and hospitalizations with one or more COVID-19-like illness (CLI)-related discharge codes *(International Classification of Diseases (ICD), Ninth and Tenth Revisions)* (eTable 2) and a molecular test for SARS-CoV-2 performed within 14 days before or up to <72 hours after the encounter during a BA.4/BA.5 predominant period of June 19 to August 20, 2022. Site-specific start dates were defined from local sequencing data when the combined prevalence of BA.4 and BA.5 was ≥50% and continued through the end of the study period (August 20, 2022). COVID-19 cases included patients with ≥1 CLI code(s) and a positive SARS-CoV-2 molecular test; controls included patients with ≥1 CLI code(s) and a negative SARS-CoV-2 molecular test.

For the comparison of severity by Omicron sublineage period, we included COVID-19 cases hospitalized during all Omicron sublineage periods (BA.1, combined BA.2/BA.2.12.1, and BA.4/BA.5 sublineage predominant periods). In this analysis, baseline characteristics of hospitalized COVID-19 case-patients, including demographics, underlying medical conditions, prior vaccination and prior infection histories, and in-hospital outcomes (hospital length of stay [LOS], intensive care unit [ICU] admission, invasive mechanical ventilation, and in-hospital death within 28 days of admission), were compared by period.

### Participants

This analysis included adults ≥18 years of age with a CLI-related medical encounter and SARS-CoV-2 molecular testing. CLI encounters include codes for acute respiratory clinical diagnoses (eg, pneumonia, respiratory failure) or COVID-19-related signs or symptoms (eg, shortness of breath, cough, fever) during a UC or ED visit or a hospital admission with ≥24 hours duration. Repeat ED/UC visits within a 24-hour period or multiple hospital admissions that occurred within a 30-day period (with respect to prior discharge) were combined into a single event with the earliest date used as the index date to determine vaccination status. One individual could contribute more than 1 event during the analysis period. Information on patients’ baseline characteristics, including demographic characteristics, underlying medical conditions, and prior SARS-CoV-2 testing results, was obtained through electronic medical records and ICD codes. Only completed encounters (ie, hospital events in which a patient was discharged or died) were included in this analysis.

### Classification of Vaccination Status

COVID-19 vaccination status was ascertained through state or local immunization information systems (IIS), electronic medical records, and claims data. Only mRNA vaccines (BNT162b2 from Pfizer-BioNTech or mRNA-1273 from Moderna) were considered in this analysis. Vaccination status was assigned using doses received prior to a medical encounter index date, defined as either the date of collection of a respiratory specimen associated with the most recent positive or negative SARS-CoV-2 test result before the medical visit or the date of the medical visit (if testing occurred only after the admission or visit date). Patients were considered unvaccinated if no mRNA vaccine doses were received before the index date, vaccinated with a primary series if 2 doses were received with the second dose ≥14 days before the index date, vaccinated with a first booster if 3 doses were received with the third dose ≥7 days before the index date, or, among patients aged ≥50 years, vaccinated with second booster if 4 doses were received with the fourth dose ≥7 days before the index date. Patients were excluded if they received a third or fourth dose before they were recommended for immunocompetent adults or received a dose with a shorter interval than recommended (ie, less than 5 months between second and third dose or less than 4 months between third and fourth dose). Patients were excluded if they received 1 mRNA vaccine dose, if they received a non-mRNA vaccine (eg, Janssen/Johnson & Johnson product), or if they had a likely immunocompromising condition, as previously defined.^17^

### COVID-19 severity classification

For hospitalized patients, additional clinical outcomes including ICU admission, in-hospital death up to 28 days after admission, hospital LOS, and receipt of invasive mechanical ventilation (IMV) were obtained through electronic medical records.

### Statistical Analysis

The association between symptomatic laboratory-confirmed SARS-CoV-2 infection at an ED/UC encounter or hospitalization and vaccination status was estimated among individuals with CLI using multivariable logistic regression to compare the odds of prior receipt of 2, 3, and 4 vaccine doses vs unvaccinated status (reference) in SARS-CoV-2–positive cases vs SARS-CoV-2–negative controls. VE was then calculated as⍰[1⍰–⍰OR]⍰×⍰100% for the ORs for 2, 3, and 4 doses vs unvaccinated, respectively, against COVID-19–associated ED/UC encounters or hospitalizations, with lower ORs suggesting more protection. To evaluate VE against more severe COVID-19, a further analysis was performed among hospitalized COVID-19 cases comparing only cases who were admitted to the ICU and/or experienced in-hospital death with hospitalized controls^18^. Two-dose, 3-dose, and 4-dose VE estimates were further stratified by time periods since most recent vaccination dose (ie, 2-dose 14-149 days, 2-dose ≥150 days, 3-dose 7-119 days, 3 dose ≥120 days, 4-dose 7-59 days, and 4-dose ≥60 days).

In addition to estimating absolute VE (ie, VE for receipt of vaccine compared to unvaccinated), ORs were also calculated to estimate relative VE (rVE) to determine the incremental benefit of receiving an additional vaccine dose when recommend. Relative VE was estimated by comparing individuals who had recently received 1 or 2 booster doses to those who were eligible for, but had not received, the respective booster dose, ie, 3 doses within the last 7-119 days vs 2 doses ≥150 days prior, and 4 doses within the last 7-119 days vs 3 doses ≥120 days prior.

ORs were estimated separately among ED/UC encounters and hospitalizations for any combination of mRNA vaccine products and stratified by age group (18–49, 50-64, and ≥65 years). Additional OR estimates were calculated restricting cases to those admitted to the ICU and/or experiencing an in-hospital death, while including all hospitalized controls.^18^ Two additional sensitivity analyses were conducted: 1) stratified by vaccine product received (BNT162b2 or mRNA-1273); and 2) restricted to patients without a prior SARS-CoV-2 infection documented in electronic medical records.

All models included covariates to adjust for age, geographic region, calendar time, and level of local SARS-CoV-2 circulation (7-day moving average of percentage of RT-PCR tests that were SARS-CoV-2–positive within the medical facility’s geographic region). Age, calendar time, and SARS-CoV-2 circulation level covariates were specified as natural cubic spline functions with knots at quartiles. For models estimating the absolute OR, cases and controls were weighted by the inverse propensity to be vaccinated (if vaccinated) or unvaccinated (if not vaccinated). For models estimating relative ORs a similar methodology was used based on patients’ inverse propensity to be 3-dose vs 2-dose vaccinated or 4-dose vs 3-dose vaccinated. Generalized boosted regression trees were used to estimate the propensity to be vaccinated based on demographics, underlying medical conditions, and facility characteristics. Separate weights were calculated for each model and were truncated at the 99th percentile of the distribution of weights. After weighting, an absolute standardized mean or proportion difference (SMD) ≤0.2 was taken to indicate a negligible difference in distributions of covariates by vaccination status. Any covariates with SMD >0.20 after weighting were also included in the model in addition to the a *priori* variables for the respective OR estimate to minimize residual confounding (eTable 3). Two-sided 95% CIs were calculated for each reported OR, with 95% CIs that excluded 1 considered statistically significant. Non-overlapping CIs were interpreted as statistically different ORs.

To describe outcomes of patients hospitalized with COVID-19 during the BA.4/BA.5 predominant period compared to earlier sublineage periods, this analysis was restricted to hospitalized case-patients during BA.1, BA.2/BA.2.12.1, and BA.4/BA.5 periods who met other inclusion criteria above. Baseline demographic, clinical, and vaccination characteristics and in-hospital outcomes were compared between BA.4/BA.5 cases and those during other sublineage periods using SMDs.

Analyses were performed using R software, version 4.1.2 (R Foundation for Statistical Computing) and SAS, version 9.4 (SAS Institute). This activity was reviewed by CDC and was conducted consistent with applicable federal law and CDC policy. (See eg, 45 C.F.R. part 46.102(l)(2), 21 C.F.R. part 56; 42 U.S.C. §241(d); 5 U.S.C. §552a; 44 U.S.C. §3501 et seq.) Detailed methods are included in eAppendix 1 and eTables 1-2.

## RESULTS

### Participants included in VE analysis

During the BA.4/BA.5 period from June 19 to August 20, 2022, there were 253,367 ED/UC encounters and 56,471 hospitalizations at VISION Network sites; 171,138 and 35,464 patients in ED/UC encounters and hospitalizations settings, respectively, were excluded from this analysis. The most common reason for exclusion was not having molecular SARS-CoV-2 testing ≤ 14 days before to <72 hours after admission (Figure 1).

**Figure 1.**
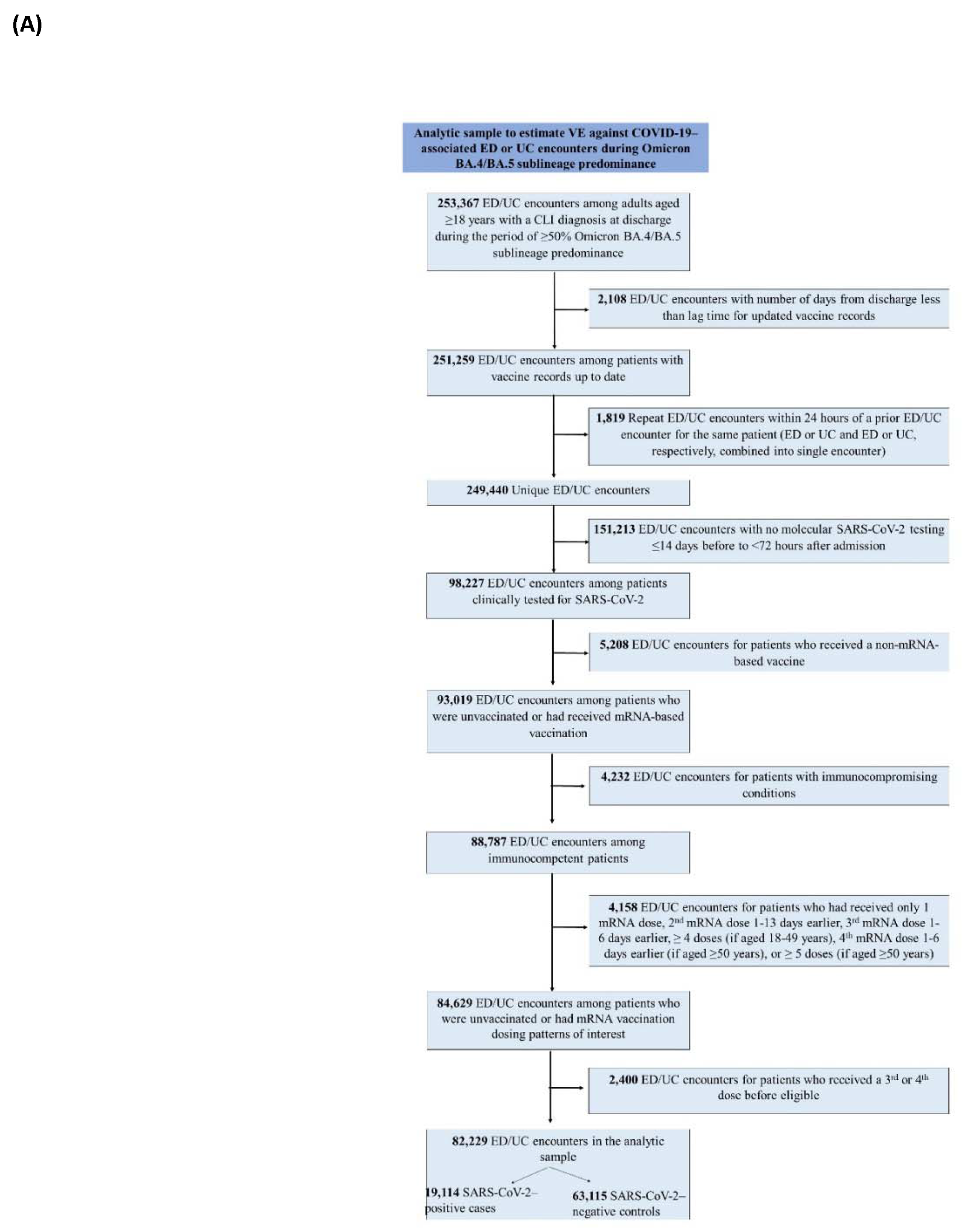
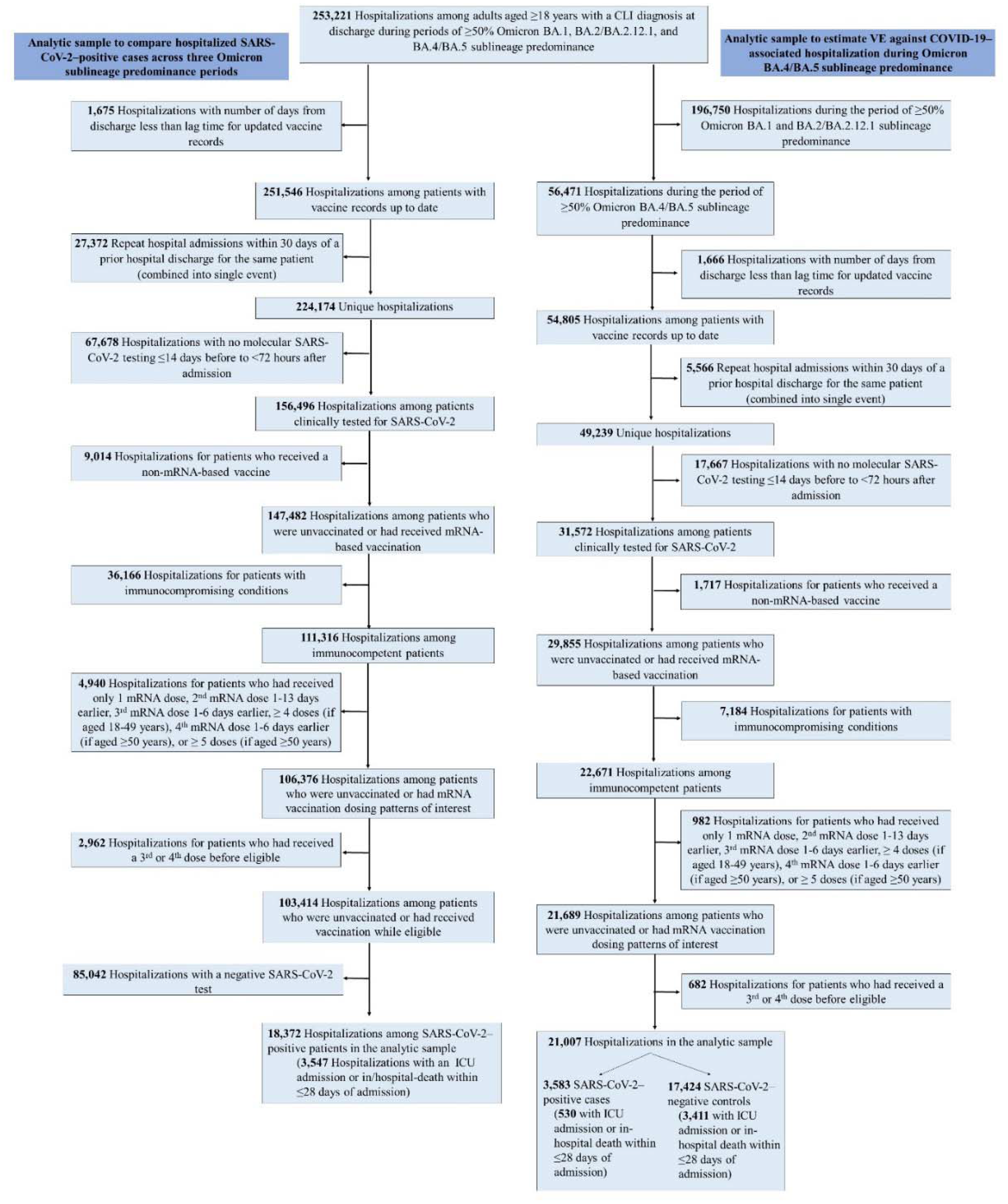
Flow Diagram for the Selection of A) Emergency Department and Urgent Care Encounters; and B) Hospitalizations Legend: CLI indicates COVID-19–like illness; ED, emergency department; ICU, intensive care unit; UC, urgent care; VE, vaccine effectiveness.

Among 82,229 ED/UC encounters included in the analysis, 19,114 encounters were cases with COVID-19 and 63,115 were controls without COVID-19 (Table 1). Cases were less likely to have received at least one booster dose compared to controls (32.3% vs 41.9%, respectively) but were similar in age to controls (median age = 50 vs 52 years). Among patients over the age of 50 years, 1,097 (11.4%) cases with ED/UC encounters had received a second booster (fourth dose) vs 6,565 (19.6%) of controls.

**Table 1.**
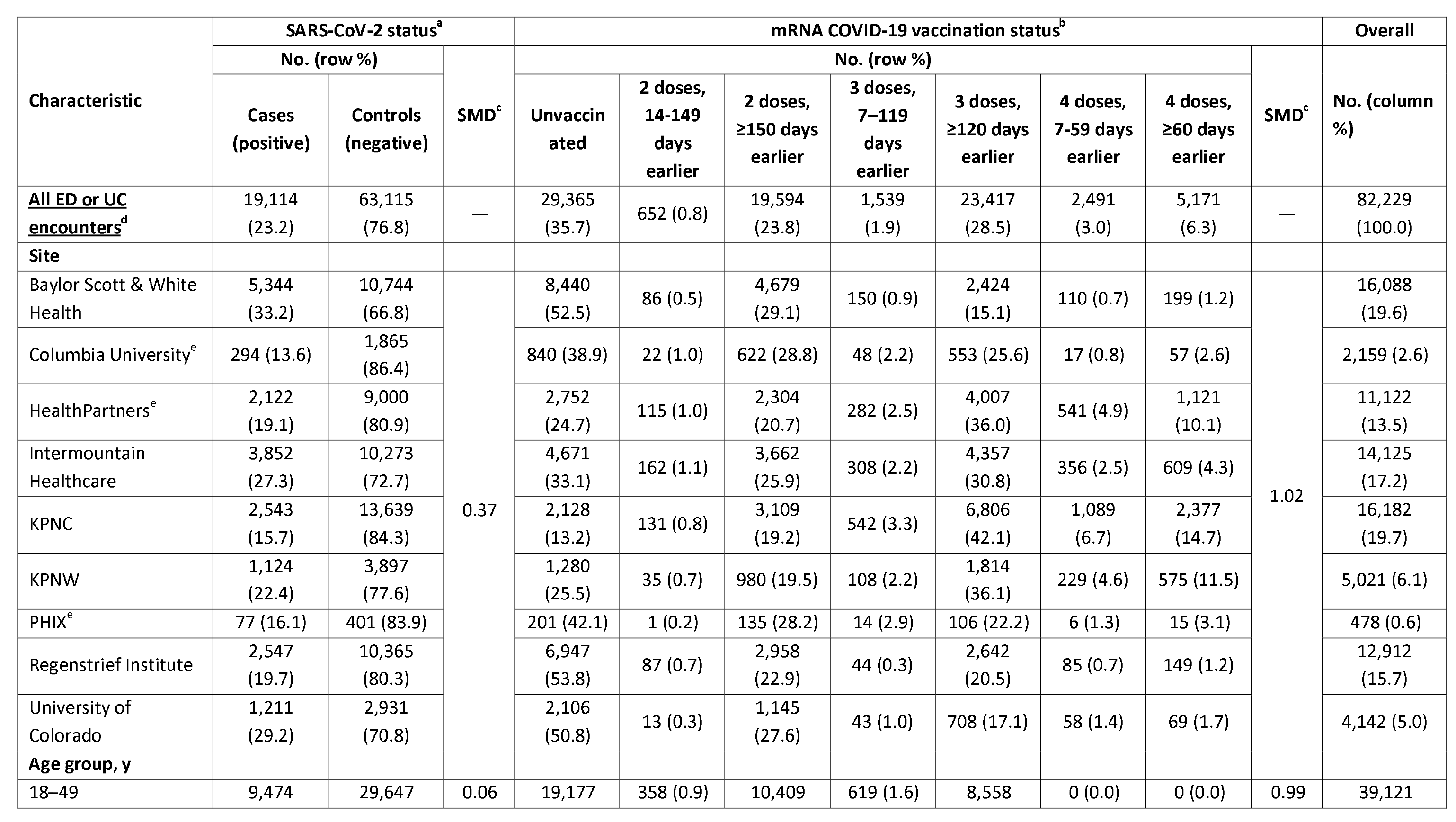
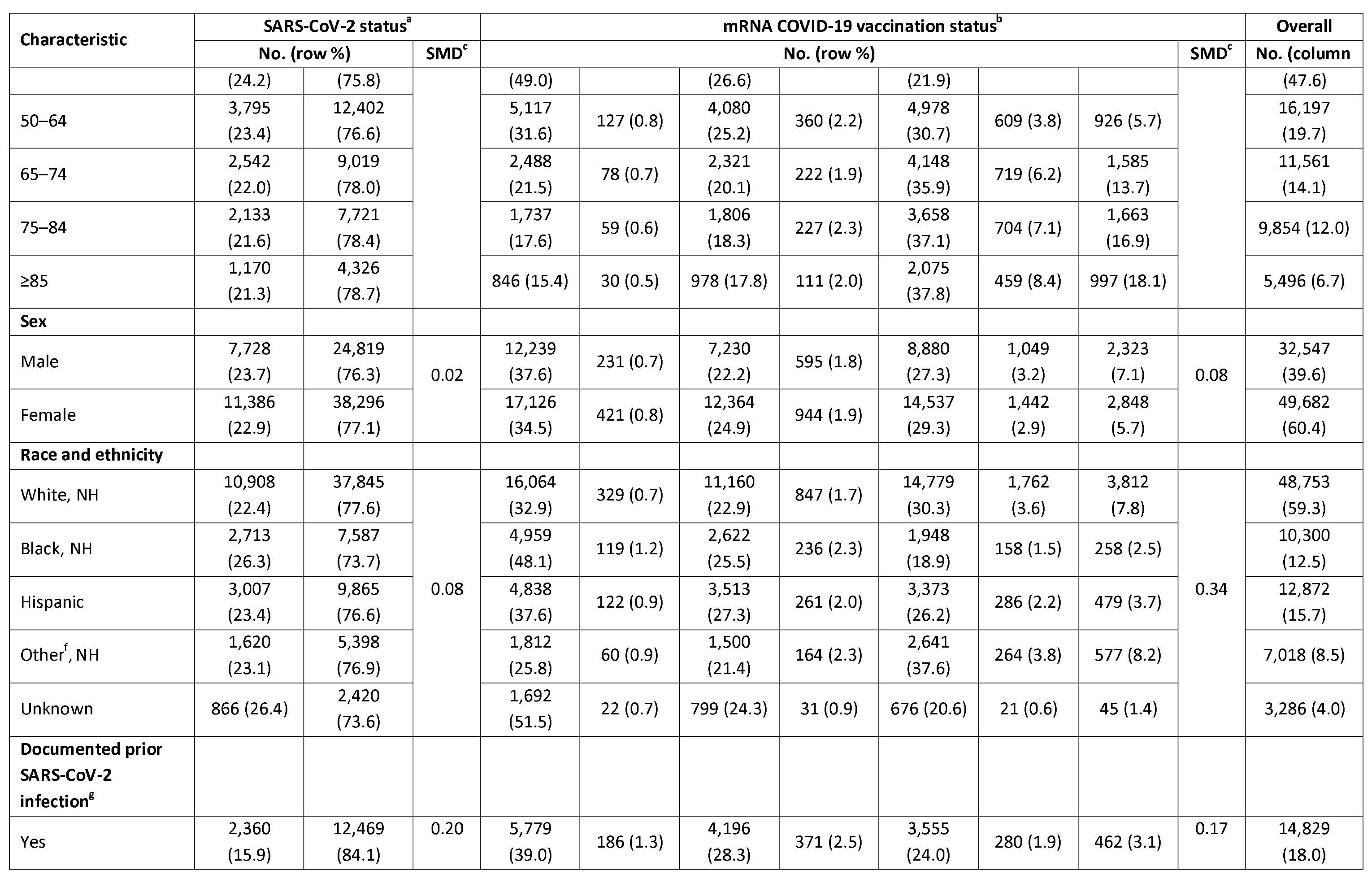
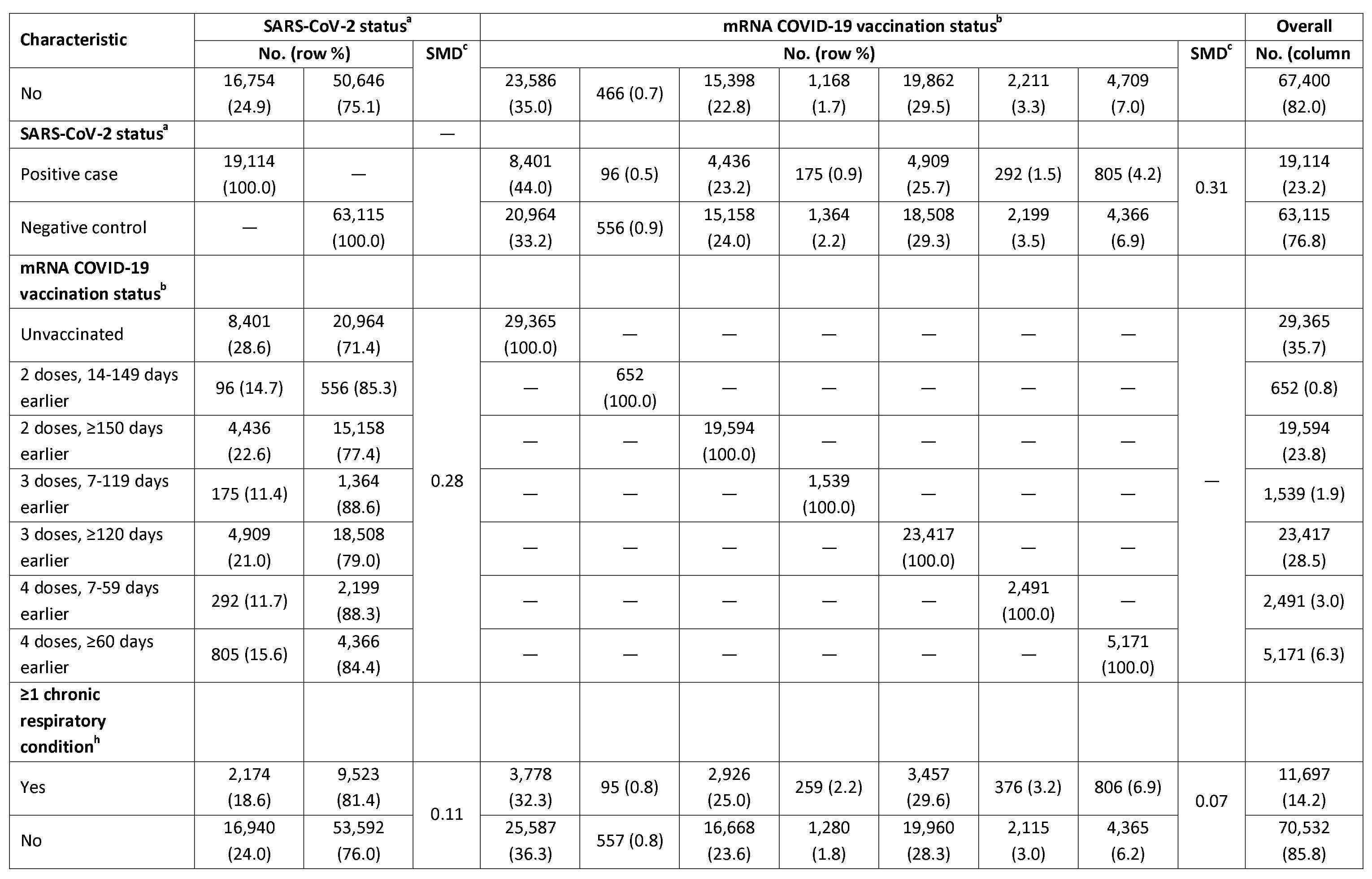
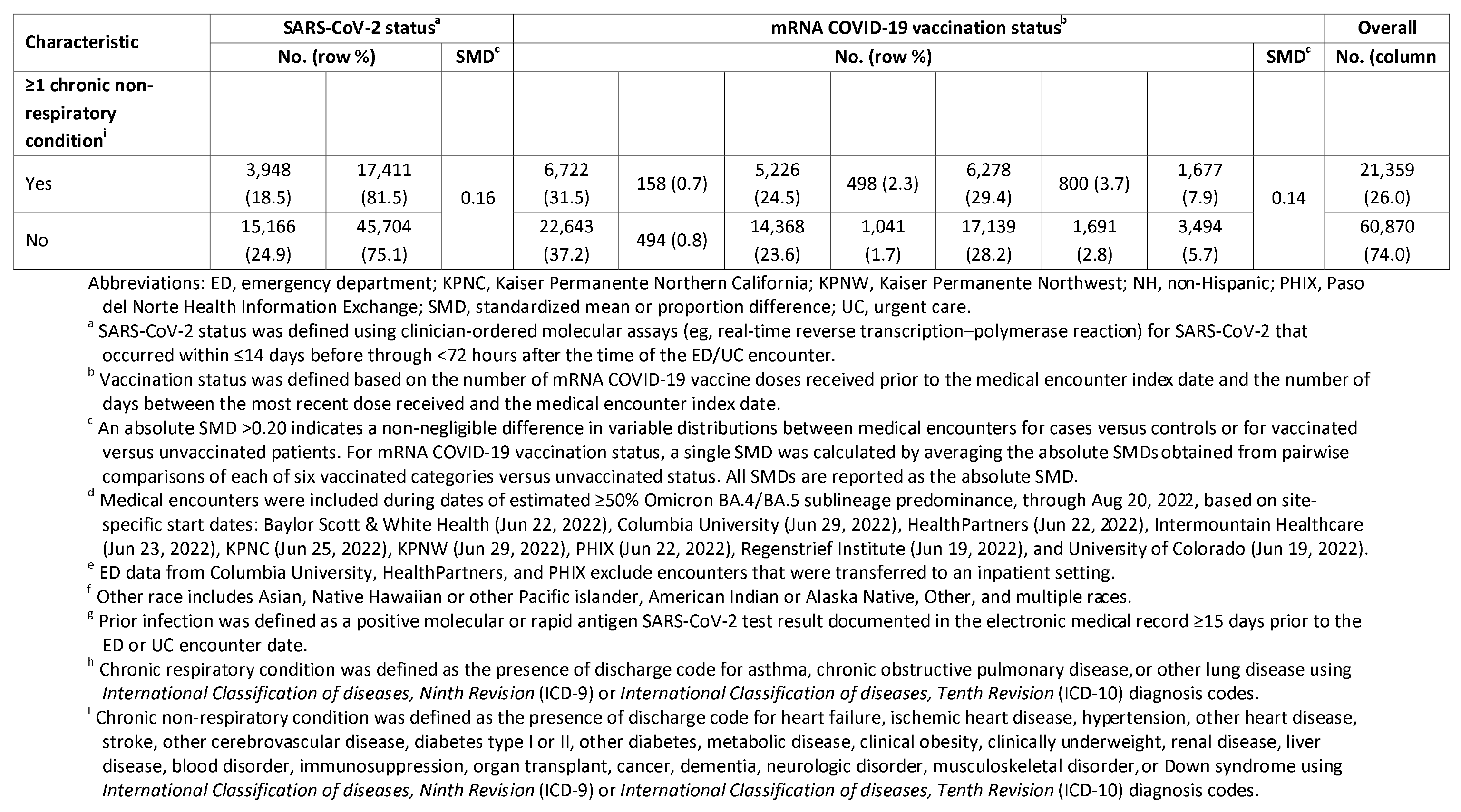
Characteristics of Emergency Department or Urgent Care Encounters Included in a Case-Control Analysis of the Association Between Symptomatic SARS-CoV-2 Infection and Prior mRNA COVID-19 Vaccination in Adults, by Case-Control Status and Vaccination Status, During a Period of Omicron BA.4/BA.5 Sublineage Predominance, VISION Network, June 19–August 20, 2022.

Among 21,007 hospitalizations included in the analysis, 3,583 were cases with COVID-19 and 17,424 were controls without COVID-19 (Table 2). Cases were less likely to have received at least one booster dose compared to controls (41.2% vs 47.1%) and more likely to be older (median age = 75 vs 70 years, respectively). Among hospitalized patients over the age of 50 years, 324 (10.3%) cases had received a second booster (fourth dose) vs 2,275 (15.8%) of controls.

**Table 2.**
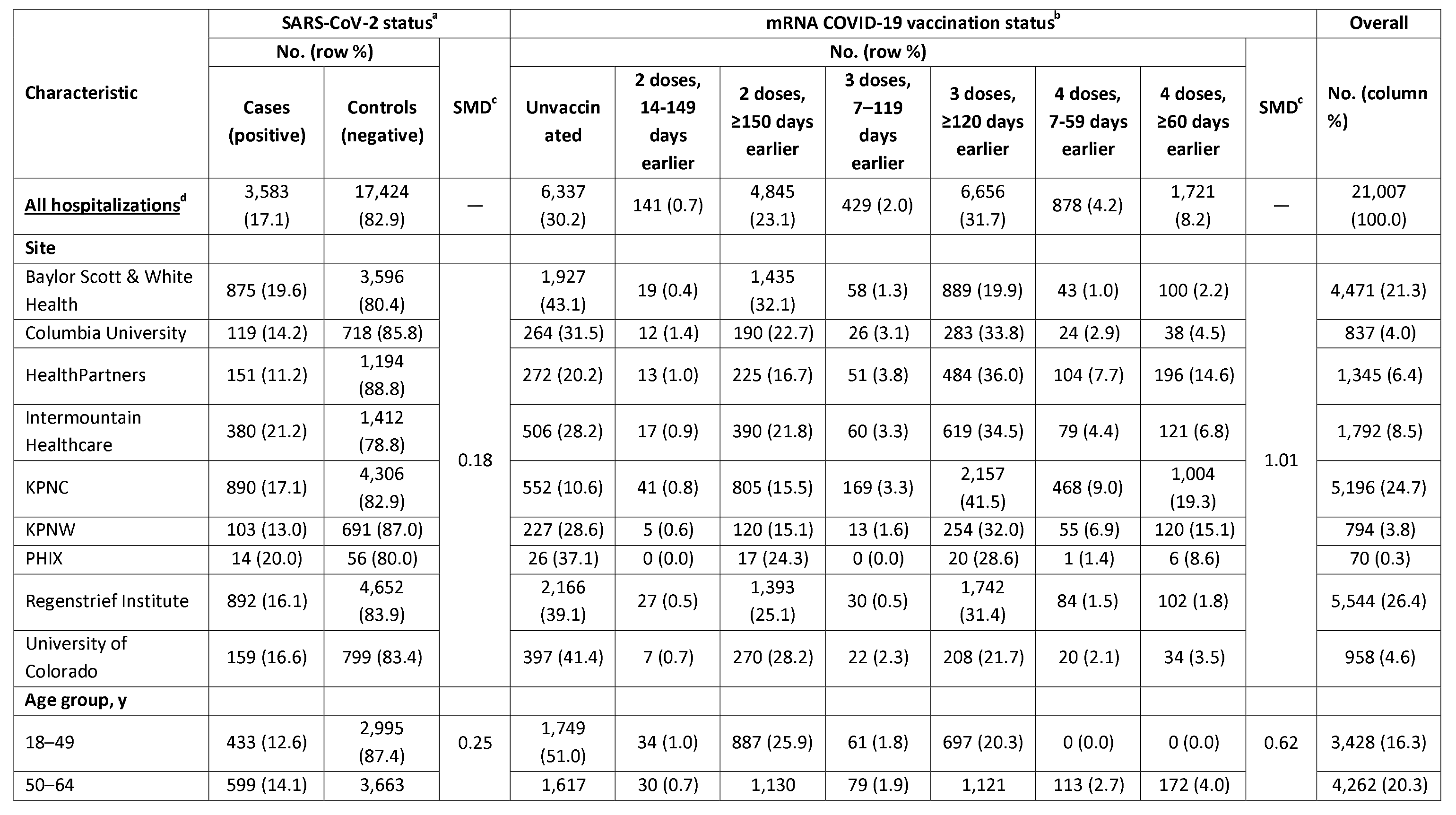
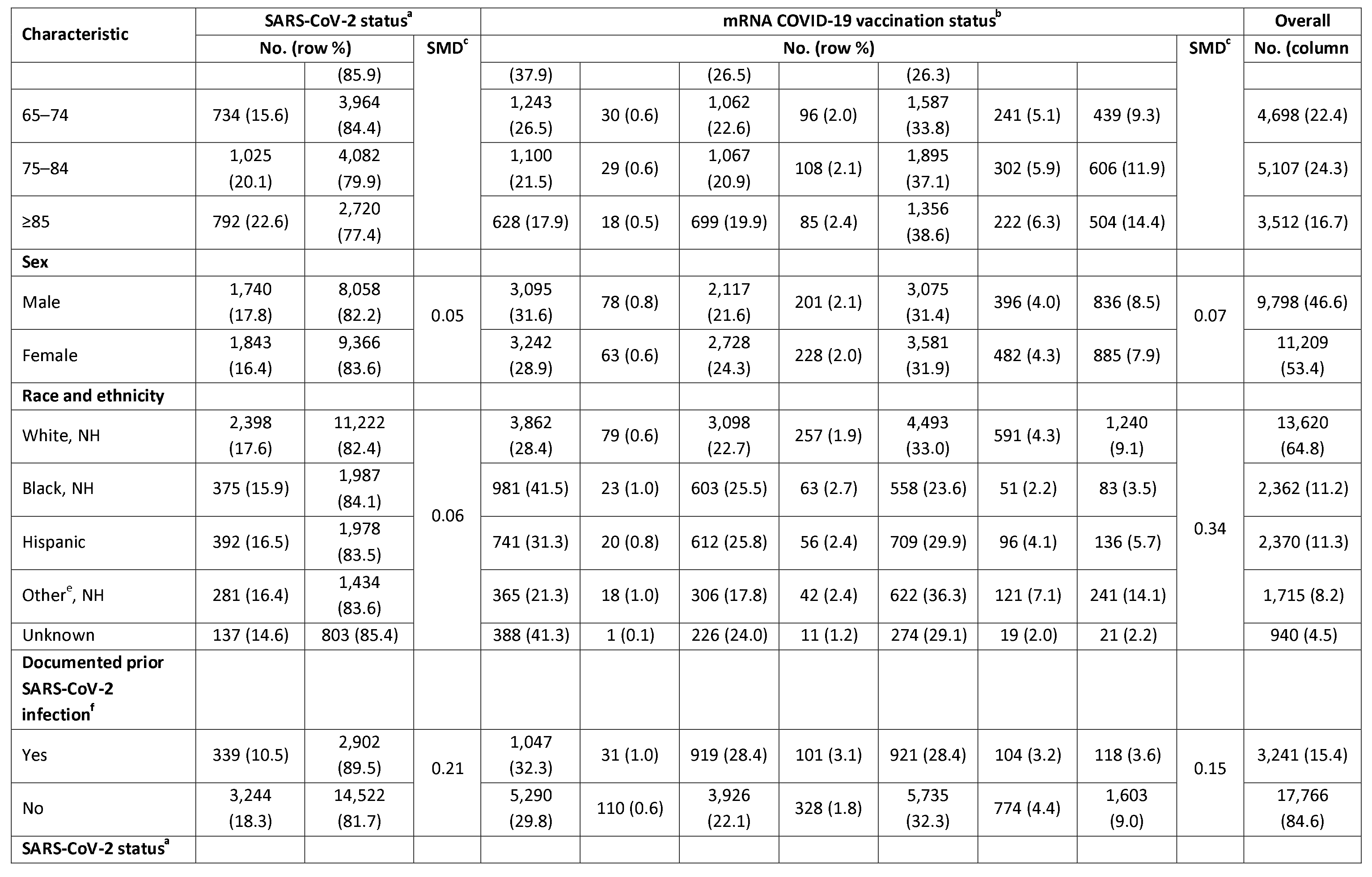
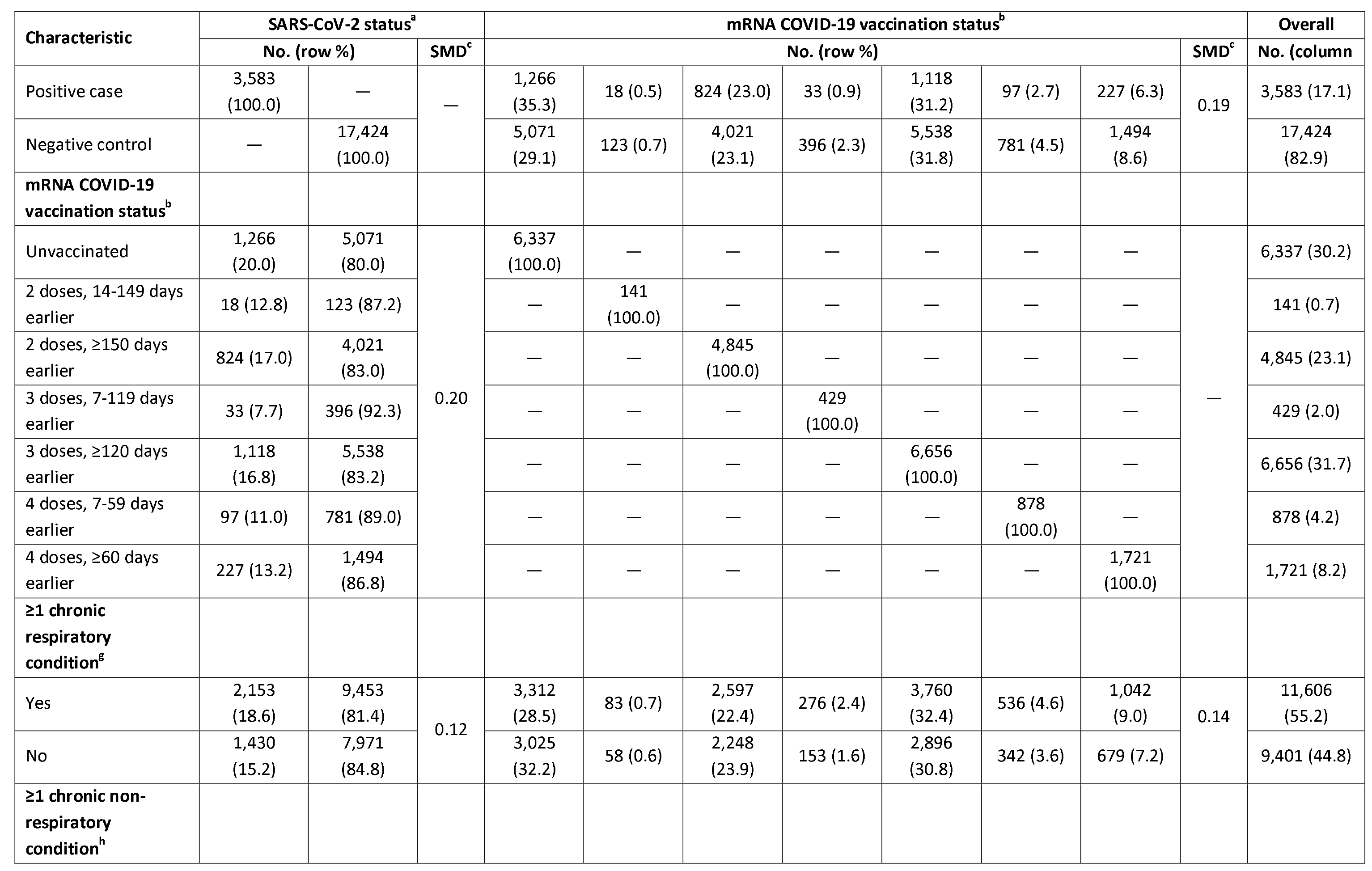
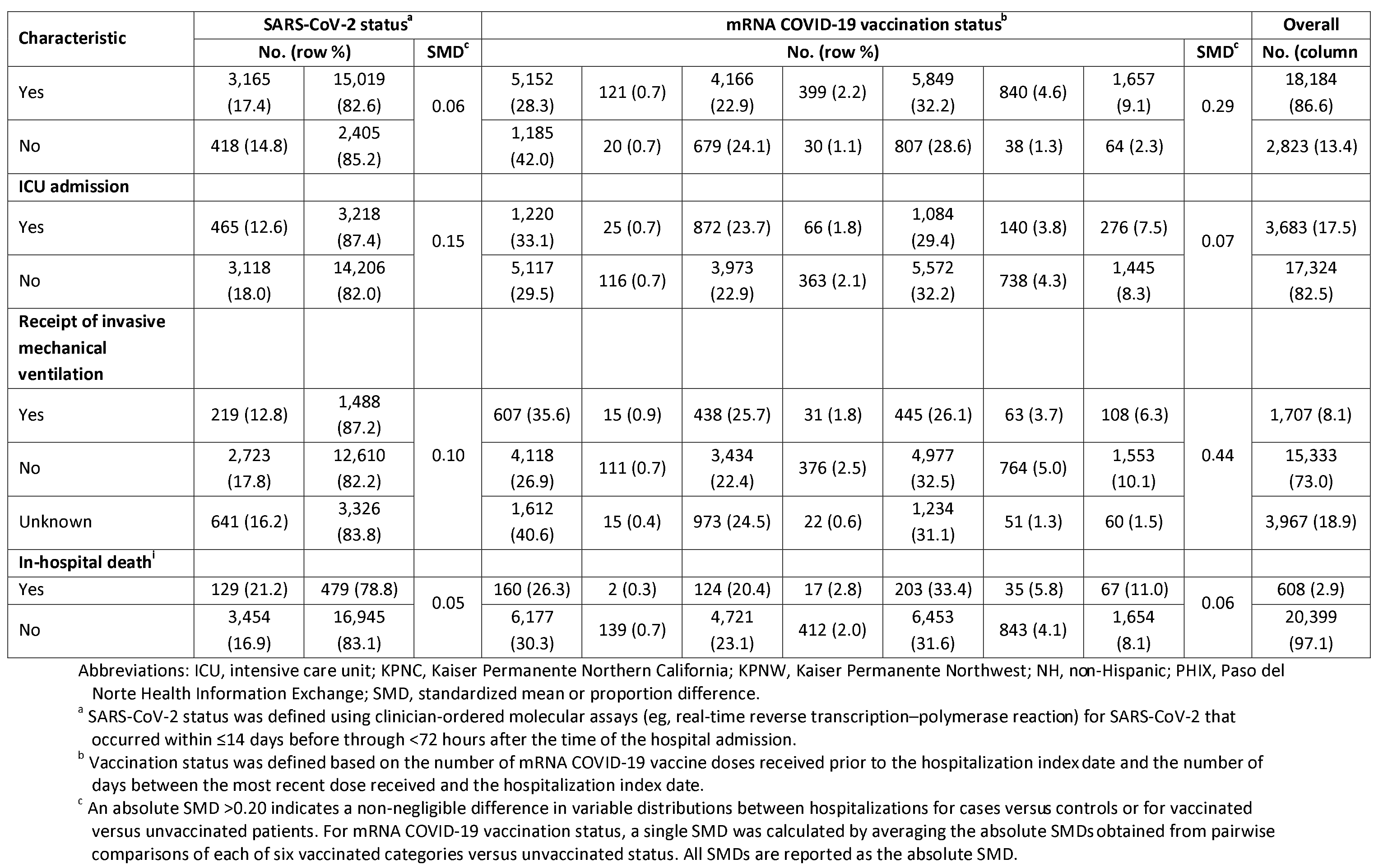
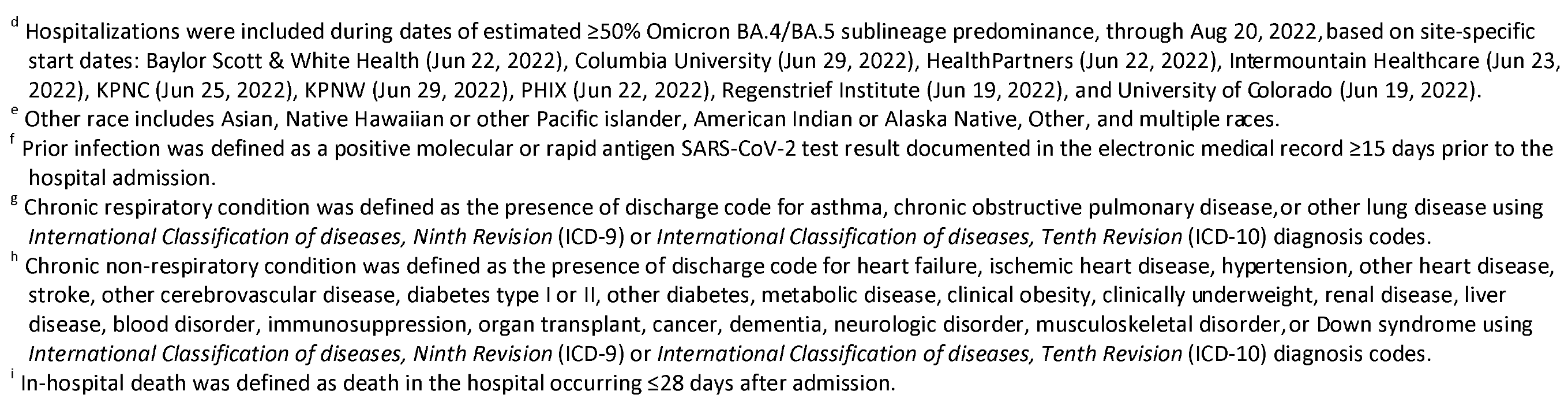
Characteristics of Hospitalizations Included in a Case-Control Analysis of the Association Between Symptomatic SARS-CoV-2 Infection and Prior mRNA COVID-19 Vaccination in Adults, by Case-Control Status and Vaccination Status, During a Period of Omicron BA.4/BA.5 Sublineage Predominance, VISION Network, June 19–August 20, 2022.

### Comparison of 2, 3, or 4 mRNA vaccine doses vs unvaccinated

Among all ED/UC encounters, the adjusted OR for prior receipt of 2 vaccine doses ≥150 days earlier (median 424 days) vs unvaccinated was 0.72 (95% CI: 0.69-0.76; estimated VE, 28% [24%-31%]) for all adults (Figure 2). The adjusted OR for a third dose 7-119 days earlier was 0.38 (95% CI: 0.32-0.46; estimated VE, 62% [54%-68%]), but the adjusted OR for the third dose ≥120 days earlier (median 228 days) was 0.68 (95% CI: 0.64-0.71; estimated VE, 32% [29%-36%]), similar to that observed with the second dose ≥150 days earlier. Among those ≥50 years eligible for a fourth dose, a fourth dose in the prior 7-59 days was associated with higher protection but associated protection also appeared to decline at ≥60 days with an adjusted OR closer to null (ie, 1).

**Figure 2.**
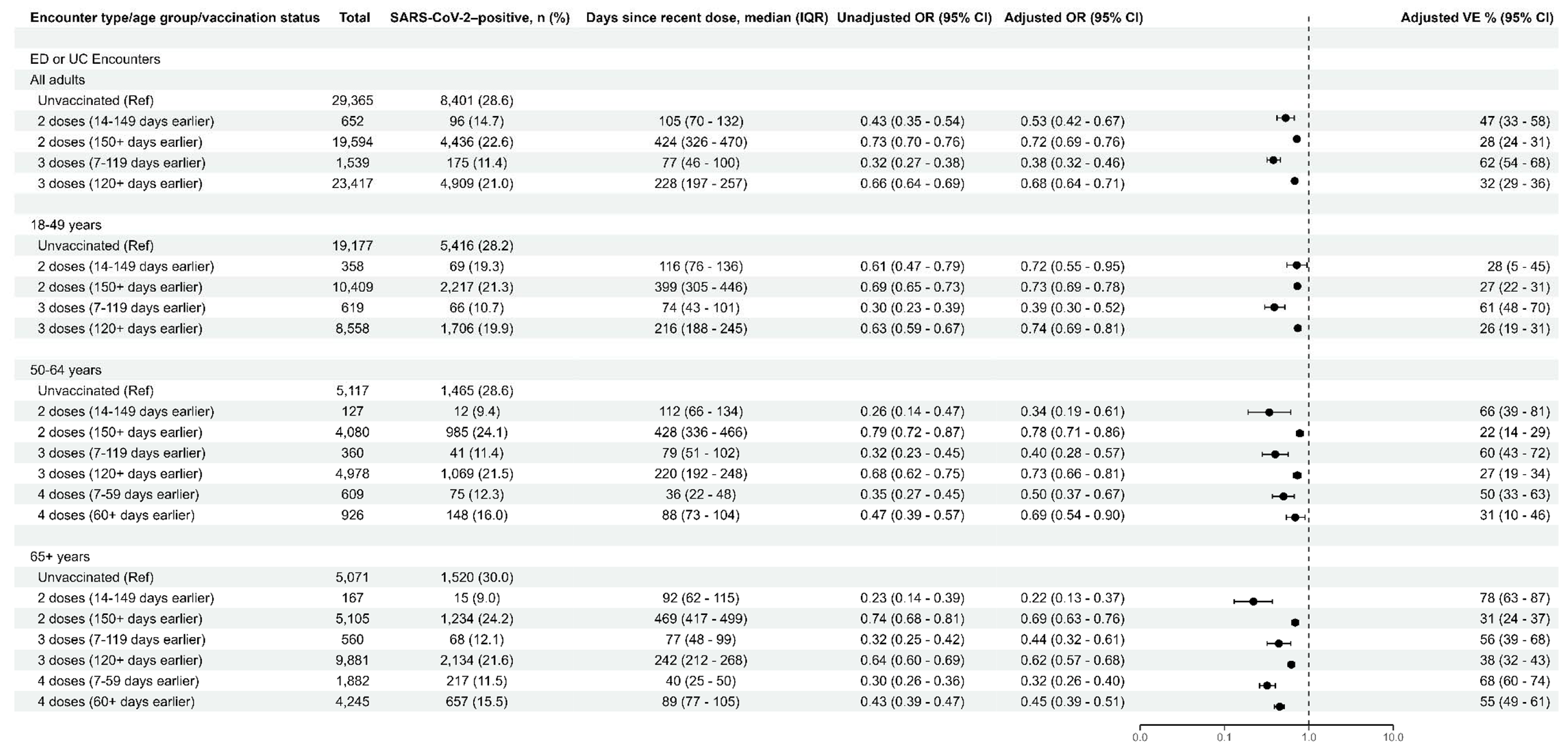
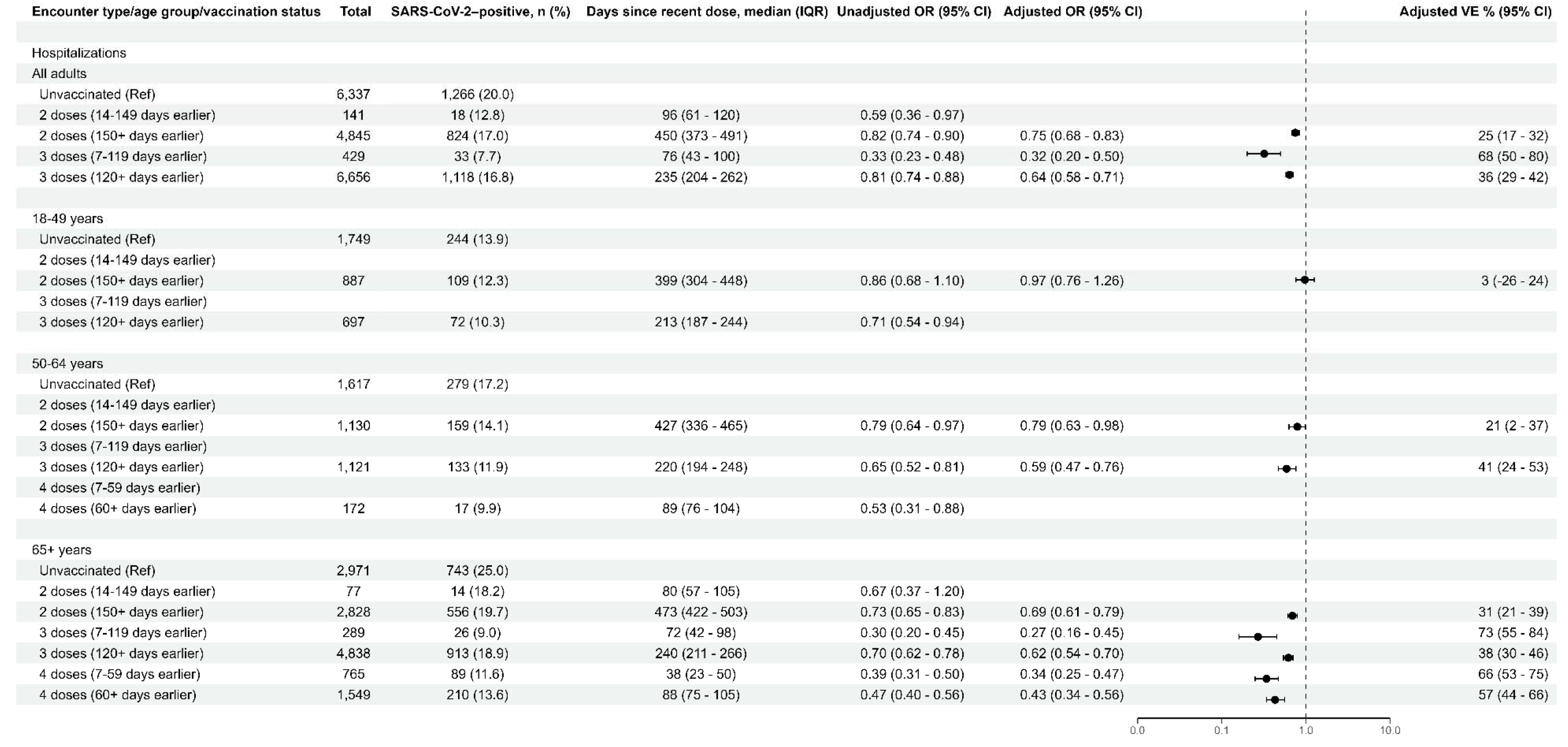
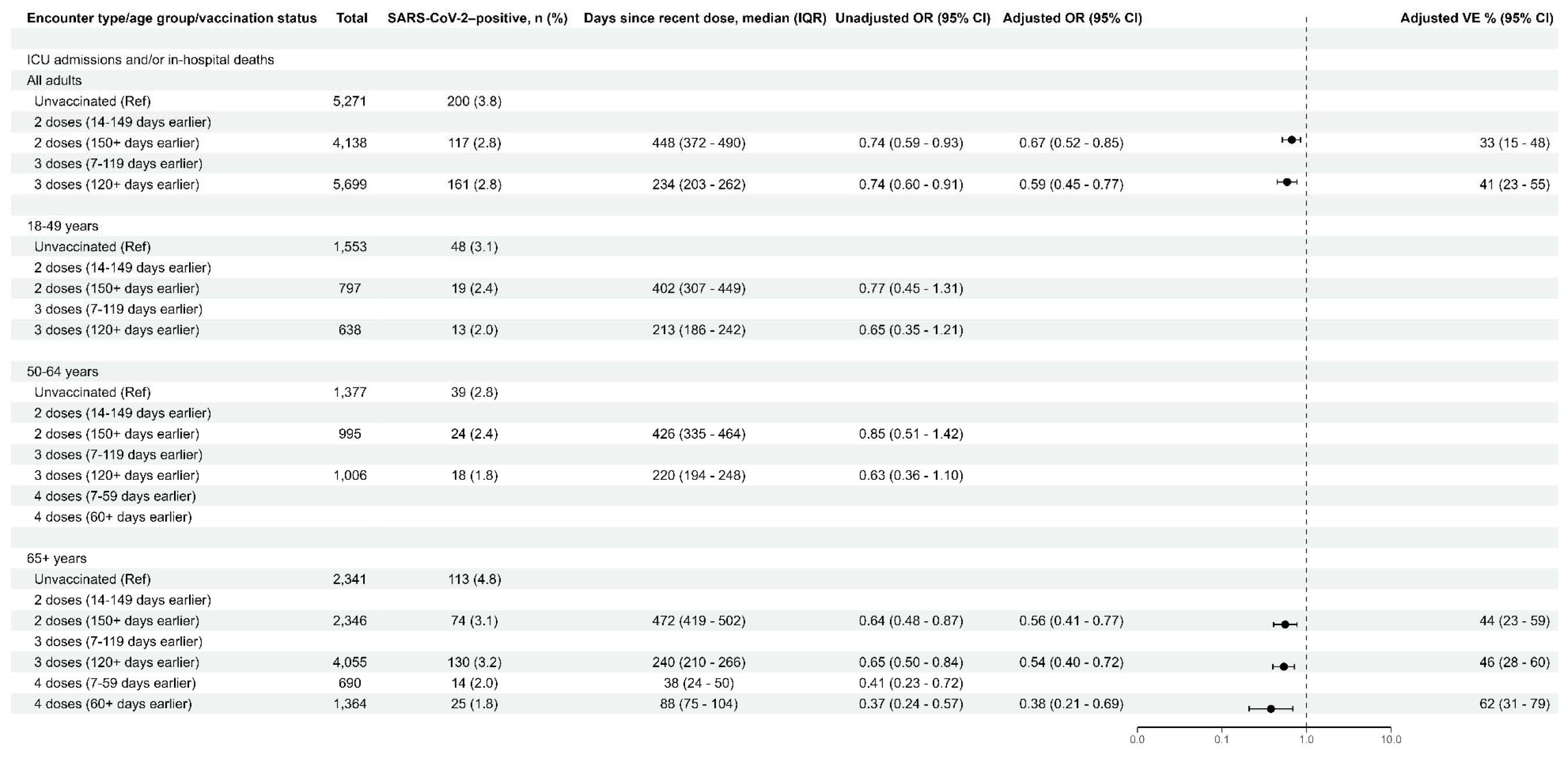
Association of COVID-19–Associated (A) Emergency Department or Urgent Care Encounters, (B) Hospitalization, and (C) Intensive Care Unit Admission and/or In-Hospital Death with Prior Vaccination with Two, Three, or Four mRNA Vaccine Doses, by Age Group, During a Period of Omicron BA.4/BA.5 Sublineage Predominance, June 19–August 20, 2022. ED or UC Encounters Hospitalizations Intensive Care Unit Admission and/or In-Hospital Death Legend: An adjusted OR <1.0 indicates that COVID-19–associated ED/UC encounter, hospitalization, or ICU admission and/or in-hospital death was associated with being unvaccinated compared with being vaccinated. ORs were adjusted for age, geographic region, calendar time (days since January 1, 2021), and local virus circulation (percentage of SARS-CoV-2–positive results from testing within the counties surrounding the facility on the date of the encounter) and weighted for inverse propensity to be vaccinated or unvaccinated (calculated separately for each OR estimate). Generalized boosted regression trees were used to estimate the propensity to be vaccinated based on the following socio-demographic, facility, and medical factors: age, sex, race, ethnicity, Medicaid status, calendar date, geographic region, local SARS-CoV-2 circulation on the day of each medical visit, urban-rural classification of facility, hospital type, number of hospital beds, chronic respiratory condition, chronic non-respiratory condition, asthma, chronic obstructive pulmonary disease, other chronic lung disease, heart failure, ischemic heart disease, hypertension, other heart disease, stroke, other cerebrovascular disease, diabetes type 1, diabetes type 2, diabetes due to underlying conditions or other specified diabetes, other metabolic disease (excluding diabetes), clinical obesity, clinical underweight, renal disease, liver disease, blood disorder, dementia, other neurological/musculoskeletal disorder, Down syndrome, and the presence of at least one prior molecular or rapid antigen SARS-CoV-2 test record documented in the electronic medical record ≥15 days before the medical encounter date (pre-vaccination, if vaccinated). Vaccine effectiveness for prevention of COVID-19–associated ED/UC encounter, hospitalization, or ICU admission and/or in-hospital death can be estimated from the adjusted ORs presented in this table with the equation: vaccine effectiveness = (1-adjusted OR) × 100%. In-hospital death was defined as death in the hospital occurring ≤28 days after admission. Adjusted ORs and VE estimates are not shown for vaccination status comparisons with confidence intervals greater than 50 percentage points around the VE estimate. Adjusted ORs and VE estimates could not be calculated for the following subgroups due to lack of model convergence: hospitalizations, 50-64 years, 2 doses (14-149 days earlier); ICU admission and/or in-hospital death, 18-49 years, 3 doses (7-119 days earlier); ICU admission and/or in-hospital death, 50-64 years, 2 doses (14-149 days earlier); and ICU admission and/or in-hospital death, 65+ years, 2 doses (14-149 days earlier). In vaccination status subgroups with <10 SARS-CoV-2–positive cases, all numbers in the row were removed because of small cell sizes. Analyses for ICU admission and/or in-hospital death included SARS-CoV-2–positive cases with ICU admission and/or in-hospital death and all SARS-CoV-2– negative hospitalized controls. CI indicates confidence interval; ED, emergency department; ICU, intensive care unit; IQR, interquartile range; OR, odds ratio; Ref, referent group; UC, urgent care; VE, vaccine effectiveness.

Among hospitalizations, the adjusted OR for prior receipt of 2 vaccine doses ≥150 days earlier vs unvaccinated was 0.75 (95% CI: 0.68-0.83; estimated VE, 25% [17%-32%]) (Figure 2). A recent third dose was associated with increased protection (adjusted OR = 0.32, 95% CI: 0.20-0.50; estimated VE, 68% [50%-80%]), but the adjusted OR was closer to 1 at ≥120 days after receipt (adjusted OR = 0.64, 95% CI: 0.58-0.71; estimated VE, 36% [29%-42%]), suggestive of waning effectiveness. Among adults ≥65 years of age, a fourth dose was associated with greater protection compared to a late third dose that was similar at 7-59 (adjusted OR = 0.34, 95% CI: 0.25-0.47; estimated VE, 66% [53%-75%]) and ≥60 days (adjusted OR = 0.43, 95% CI: 0.34-0.56; estimated VE, 57% [44%-66%]) after receipt of the fourth dose. Adjusted ORs were similar or lower (indicating higher associated protection) against COVID-19-associated ICU admission and/or in-hospital death and among those ≥65 years of age who received a fourth dose (Figure 2c).

In sensitivity analyses, findings were generally similar and with overlapping confidence intervals between BNT162b2 and mRNA-1273 recipients (eFigure 1-3) and when restricted to patients without a laboratory-confirmed history of prior SARS-CoV-2 infection (eFigure 4-6).

### Comparison of 3 vs 4 doses and 2 vs 3 doses

Receipt of a third dose within the previous 7-119 days was associated with greater protection compared to completing two doses ≥150 days earlier among all adult patients’ ED/UC encounters (adjusted OR for 3 recent doses vs 2 distant doses = 0.51, 95% CI: 0.42-0.61; estimated rVE, 49% [39%-58%]) and hospitalizations (adjusted OR = 0.43, 95% CI: 0.28-0.65; estimated rVE, 57% [35%-72%]) (eTables 4-5). Likewise, in older adults ≥65 years of age, a recent fourth dose was associated with greater protection than a distant third dose among patients’ ED/UC encounters (adjusted OR for 4 vs 3 doses = 0.65, 95% CI: 0.59-0.72; estimated rVE, 35% [28%-41%]) and hospitalizations (OR=0.63, 95% CI: 0.54-0.75; estimated VE, 37% [25%-46%]). There was not enough statistical power to calculate precise estimates against ICU admission or in-hospital death.

### Differences in epidemiology and outcomes of hospitalized COVID-19 cases by sublineage period

In addition to the 3,547 hospitalized COVID-19 cases included during the BA.4/BA.5 period, 12,127 hospitalized cases during the BA.1 period and 2,698 cases during the BA.2/BA.2.12.1 period were included (Table 3). Baseline characteristics and outcomes of hospitalized patients during the BA.4/BA.5 and BA.2/BA.2.12.1 periods were similar. Compared to the earlier BA.1 period, however, patients hospitalized with COVID-19 during the BA.4/BA.5 period were older (median age 75 vs 67 years; SMD 0.36) and more likely to be vaccinated (64.4% vs 35.9%; SMD across vaccination exposure groups 1.19). The severity of cases during the BA.4/BA.5 period was lower compared to the BA.1 period, with ICU admission occurring in 12.9% vs 17.6%, respectively (SMD 0.13), in-hospital death in 3.6% vs 8.4% (SMD 0.21), and shorter LOS (median 4 vs 5 days; SMD 0.31).

**Table 3.**
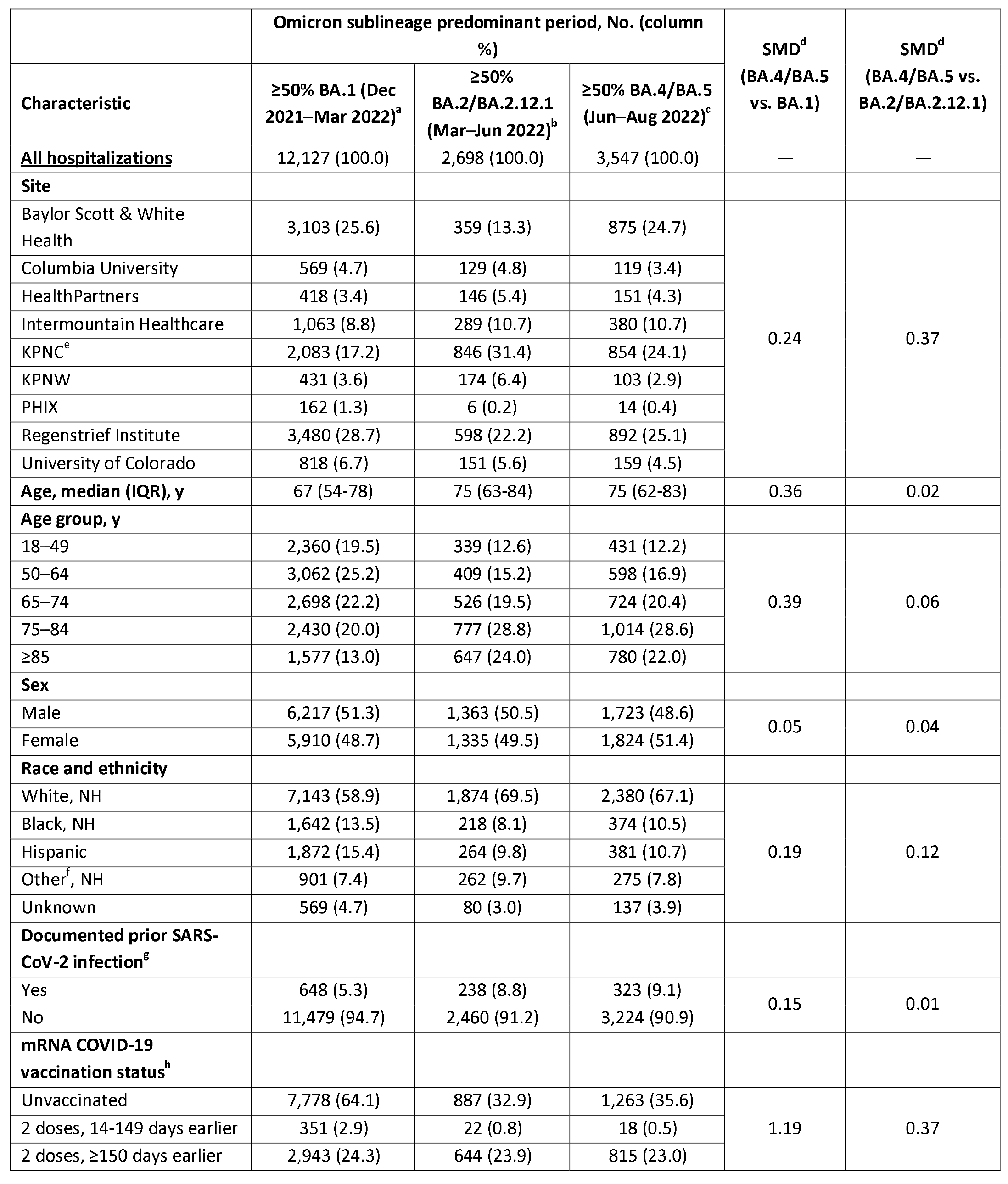
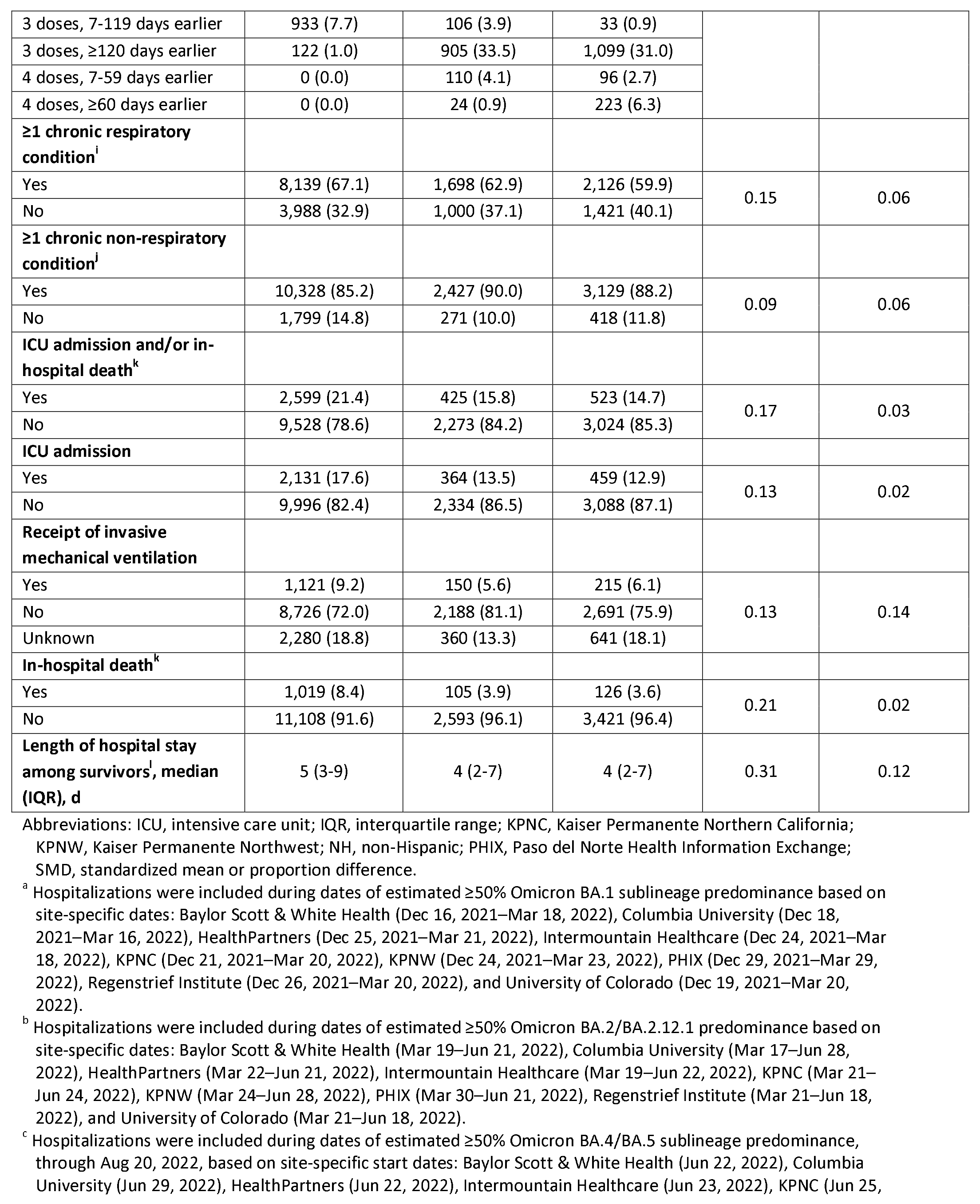
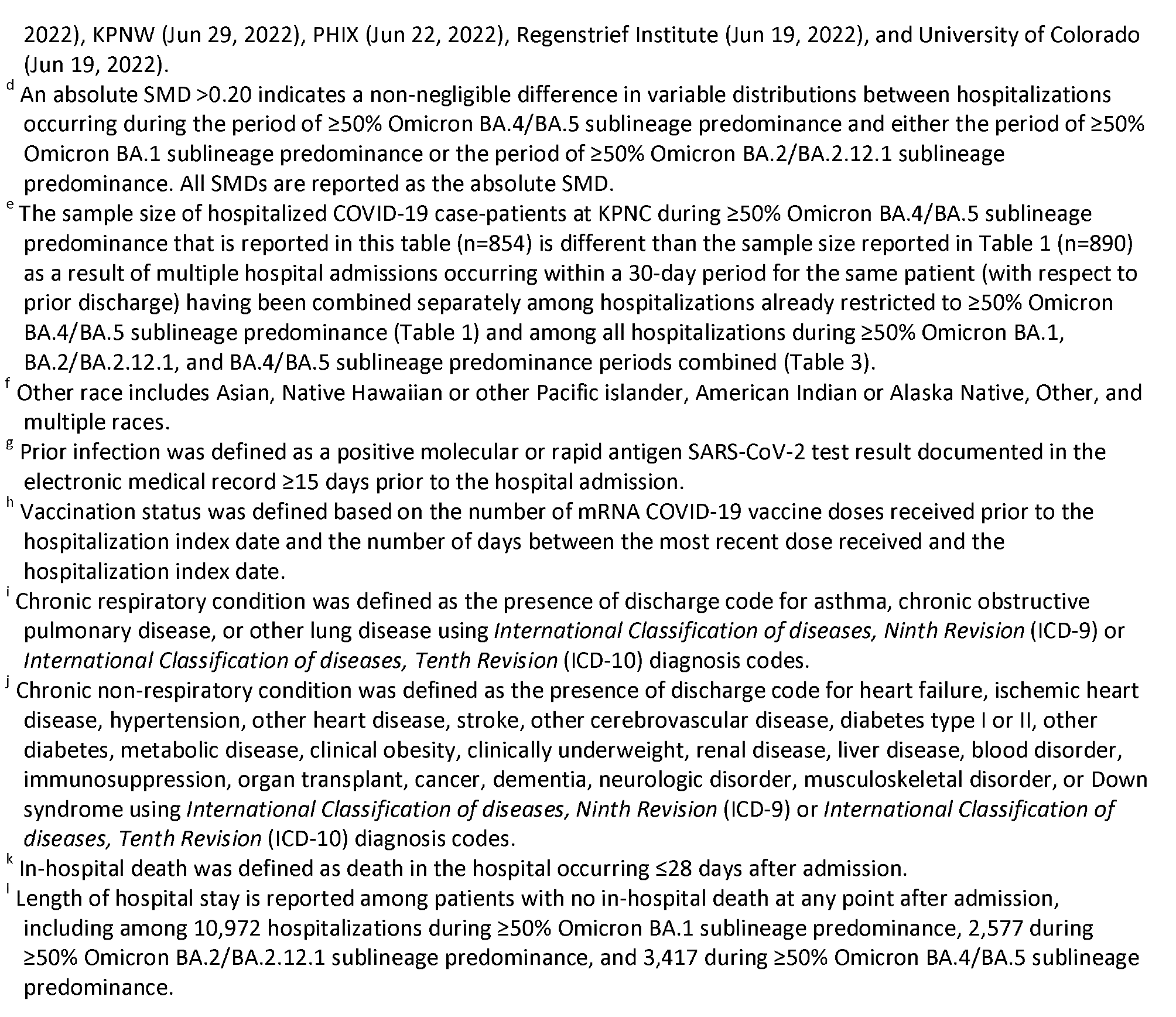
Characteristics of Hospitalized COVID-19 Case-Patients by Omicron Sublineage Predominant Period, VISION Network, December 16, 2021–August 20, 2022.

## DISCUSSION

In a multi-state analysis during Omicron BA.4 and BA.5 predominant circulation, first-generation COVID-19 vaccines remained effective against COVID-19, including for COVID-19-associated hospitalization and ICU admission or in-hospital death. However, associated protection declined within several months of the most recent vaccine dose suggesting that closeness to the most recent immunizing event impacted vaccine protection during Omicron BA.4/BA.5 variant predominance. For hospitalization, VE of 3 doses for all adults and 4 doses for adults aged ≥50 years using an unvaccinated reference group was similar to that reported during BA.2/BA.2.12.1 predominance.^19^ In addition, changes in the epidemiology of hospitalized COVID-19 cases were observed; 64% of patients hospitalized with COVID-19 during the BA.4/BA.5 predominant period had received at least a primary vaccine series vs 36% hospitalized during the earlier BA.1 predominant period. Cases hospitalized during the recent BA.4/BA.5 predominant period tended to be less severe compared with the earlier BA.1 period despite being older by 8 years on average. With authorization of updated bivalent mRNA booster vaccines, these findings provide an important baseline for future VE analyses.

Estimated VE was similar across outcomes, contradicting many past VE studies, including previous studies from VISION, which have tended to show higher vaccine-associated protection for more severe outcomes. This could be due to changes in baseline population immunity (eg, most adults now have evidence of prior infection), changes in behavior (eg, decreased use of social distancing and masks during recent months), or residual confounding.^3,4,20-22^ Across all outcomes, estimated VE in this analysis was lower than VE when the Delta variant and BA.1 sublineage of the Omicron variant predominated. However, the relative contribution of immune evasion from newer variants vs other factors such as influence of prior infections on VE is unclear and VE has been observed to wane over several months with first-generation vaccines.^24^ A recent correspondence from South Africa found that VE against COVID-19-associated hospitalization after receipt of 2 or 3 doses of BNT162b2 waned substantially within several months of vaccine receipt during BA.5 predominant circulation, which is similar to findings in this study.^26^ While this analysis did not find waning after the 4^th^ dose, median time from 4^th^ dose to the included encounter in our analysis was <3 months (compared to approximately 8 months post-3^rd^ dose), so waning may become evident with increased follow-up time. Approved bivalent booster doses, which include the ancestral strain as well as an additional component targeting BA.4/BA.5 viruses, might provide increased protection against currently circulating variants, although this will require future investigation.^25^

The finding of less severe disease during BA.4/BA.5 predominance compared with earlier Omicron sublineage periods has important implications for interpretation of VE over time. Between December 2021 and February 2022, prevalence of infection-induced antibodies among clinical samples tested at commercial laboratories increased from 33.5% to 57.7%, indicating widespread infection-induced immunity by the end of the BA.1 predominant period.^8^ VE estimates during BA.4/BA.5 predominance should therefore be interpreted in the context of population immunity due to prior infection; measured VE is likely blunted by high infection-induced immunity in the unvaccinated and under-vaccinated comparison groups.

Although BA.5 represented over 80% of sequenced isolates included in genomic surveillance-based estimates of variant proportions during September 2022, the original BA.4 and BA.5 sublineages have identical spike proteins, indicating that these VE estimates likely hold for both sublineages.^23^ The implications of these findings on potential vaccine protection for other emerging sublineages, such as BA.4.6, which carries an additional mutation in the spike protein, are unclear.

### Limitations

This analysis is subject to several limitations. First, patient samples were not available for genomic characterization directly. Local prevalence estimates of BA.4/BA.5, combined with date of testing, were used ecologically to determine inclusion in the analysis periods; however, as estimates of VE during BA.2/BA.2.12.1 predominance are similar to this analysis, misclassifying some early cases as BA.4/BA.5 should not have impacted VE estimates substantially. Second, because prior infection was likely under-ascertained, the primary analysis included all individuals, regardless of documented prior infection or time since documented prior infection, which may have biased results towards the null if prior infection is associated with some protection against re-infection or attenuation of severity if re-infected. Third, although inverse propensity to be vaccinated weights were used to balance vaccinated and unvaccinated medical encounters, residual confounding in VE estimates due to other factors is possible. Finally, this analysis combined estimated VE against ICU admission and death, which may have obscured differences in VE for these individual outcomes.

## CONCLUSIONS

Among immunocompetent adults, compared to unvaccinated adults, the estimated VE of recently received third or fourth doses of an mRNA vaccine against ED/UC visits, hospitalization, and ICU/death was higher compared to 2 doses, but waned during BA.4/BA.5 variant predominance. Hospitalized cases were less likely to be admitted to the ICU or experience in-hospital death and had shorter length of admission during BA.4/BA.5 predominance compared to earlier Omicron sublineage periods.

## Data Availability

Dissemination to participants and related patient and public communities: The individual level dataset from this study is held securely in limited deidentified form at the US Centers for Disease Control and Prevention. Data sharing agreements between CDC and data providers prohibit CDC from making this dataset publicly available. CDC will share aggregate study data once study objectives are complete, consistent with data use agreements with partner institutions.

## ARTICLE INFORMATION

### Conflict of Interest Disclosures

During the conduct of the study, all Westat- and Kaiser Permanente Northern California Division of Research-affiliated authors reported receiving contractual support from the CDC during the conduct of the study via payments made to respective institutions. Additionally, all authors affiliated with Baylor Scott & White Health, Children’s Minnesota Columbia University Irving Medical Center, HealthPartners Institute, Intermountain Healthcare, Kaiser Permanente Northwest, Paso del Norte Health Information Exchange, Regenstrief Institute, and University of Colorado receiving contractual support from the CDC during the conduct of the study, via a subcontract from Westat, Inc. to their institution. Outside the submitted work in the past 36 months, the following disclosures were reported: Dr Naleway reported receiving grants from Pfizer and Vir Biotechnology; Dr Klein reported receiving grants from Pfizer, Merck, GlaxoSmithKline, and Sanofi Pasteur; Dr Gaglani reported receiving grants directly from CDC and from CDC via subcontracts from Abt Associates and Vanderbilt University Medical Center to her institution; Dr Irving reported receiving grants from the CDC to her institution; Dr Dixon reported receiving grants from CDC, NIH, AHRQ, and the U.S. Department of Veterans Affairs, personal fees from Elsevier and Springer Nature, consulting fees from Merck and Co; Dr. McEvoy reported receiving grants AztraZeneca; Dr Rao reported receiving grants from GSK; and Dr Uhlemann reported receiving grants from Merck. Dr Uhlemann and Dr Annavajhala also reported receiving grants from NIH/NIDA during the conduct of the study. No other disclosures were reported.

### Additional Contributions

We would like to acknowledge the contributions of the VISION Network; Westat: Elizabeth Bassett, BA; Bria Berry, MPH; Rebecca Birch, MPH; Kevin Cheng, BS; Sumathi Croos, BA; Jonathan Davis, PhD; Maria Demarco, PhD; Rebecca Fink, MPH; Carly Hallowell, MPH; Nina Hamburg, MBA; Alex Hughes, PhD; Jean Keller, MS; Salome Kiduko, MPH; Lindsey Kirshner, MPH; Magdalene Kish, BS; Victoria Lazariu, PhD; Yong Lee, BSEE; Vanessa Masick, MS; Thomas Mienk, MPA; Patrick Mitchell, ScD; Jean Opsomer, PhD; Weijia Ren, PhD; John Riddles, PhD; Elizabeth Rowley, DrPH; Anna Rukhlya; MA, Kristin Schrader, MA; Patricia Shifflett, MS; Brenda Sun, MS; Zachary Weber, PhD; and Yan Zhuang, PhD; Baylor Scott & White Health: Deepika Konatham, BS; I-Chia Liao, MPH; Deborah Hendricks; Jason Ettlinger, MA; Joel Blais, BTh; Elisa Priest, DrPH; Michael Smith, BS; Spencer Rose, BS; Natalie Settele, PA; Jennifer Thomas, MS; Muralidhar Jatla, MD; Madhava Beeram, MD; Javed Butler, MD; and Alejandro Arroliga, MD; School of Medicine, University of Colorado Anschutz Medical Campus, Health Data Compass: David Mayer, BS; Bryant Doyle; Briana Kille, PhD; and Catia Chavez, MPH; Regenstrief Institute: Ashley Wiensch, MPH; and Amy Hancock, MPA; Center for Health Research, Kaiser Permanente Northwest: Padma Dandamudi, MPH; HealthPartners Institute: Inih Essien, OD; Sunita Thapa, MPH; and Sheryl Kane, BS; Intermountain Healthcare: Bert Lopansri, MD.

## Notes

### Funding Statement

Funding: This study was funded by the Centers for Disease Control and Prevention though contract 75D30120C07986 to Westat, Inc. and contract 75D30120C07765 to Kaiser Foundation Hospitals. Study sponsors placed no limitations on publication nor required confidentiality in reporting of results

### Author Declarations

IRB of Westat, Inc gave ethical approval for this work

## REFERENCES

1. Steele MK, Couture A, Reed C, et al. Estimated Number of COVID-19 Infections, Hospitalizations, and Deaths Prevented Among Vaccinated Persons in the US, December 2020 to September 2021. JAMA Netw Open. 07 01 2022;5(7):e2220385. doi:10.1001/jamanetworkopen.2022.20385

2. Wang L, Kainulainen MH, Jiang N, et al. Differential neutralization and inhibition of SARS-CoV-2 variants by antibodies elicited by COVID-19 mRNA vaccines. Nat Commun. 07 27 2022;13(1):4350. doi:10.1038/s41467-022-31929-6

3. Lauring AS, Tenforde MW, Chappell JD, et al. Clinical severity of, and effectiveness of mRNA vaccines against, covid-19 from omicron, delta, and alpha SARS-CoV-2 variants in the United States: prospective observational study. BMJ. 03 09 2022;376:e069761. doi:10.1136/bmj-2021-069761

4. Accorsi EK, Britton A, Fleming-Dutra KE, et al. Association Between 3 Doses of mRNA COVID-19 Vaccine and Symptomatic Infection Caused by the SARS-CoV-2 Omicron and Delta Variants. JAMA. 02 15 2022;327(7):639–651. doi:10.1001/jama.2022.0470

5. Hachmann NP, Miller J, Collier AY, et al. Neutralization Escape by SARS-CoV-2 Omicron Subvariants BA.2.12.1, BA.4, and BA.5. N Engl J Med. 07 07 2022;387(1):86–88. doi:10.1056/NEJMc2206576

6. Wang Q, Guo Y, Iketani S, et al. Antibody evasion by SARS-CoV-2 Omicron subvariants BA.2.12.1, BA.4 and BA.5. Nature. 08 2022;608(7923):603–608. doi:10.1038/s41586-022-05053-w

7. Centers for Disease Control and Prevention. CDC COVID Data Tracker. Available at: https://covid.cdc.gov/covid-data-tracker/#cases_casesper100klast7days. Accessed on: 4 May 2022.

8. Clarke KEN, Jones JM, Deng Y, et al. Seroprevalence of Infection-Induced SARS-CoV-2 Antibodies - United States, September 2021-February 2022. MMWR Morb Mortal Wkly Rep. Apr 29 2022;71(17):606–608. doi:10.15585/mmwr.mm7117e3

9. Ferdinands JM, Rao S, Dixon BE, et al. Waning 2-Dose and 3-Dose Effectiveness of mRNA Vaccines Against COVID-19-Associated Emergency Department and Urgent Care Encounters and Hospitalizations Among Adults During Periods of Delta and Omicron Variant Predominance - VISION Network, 10 States, August 2021-January 2022. MMWR Morb Mortal Wkly Rep. Feb 18 2022;71(7):255–263. doi:10.15585/mmwr.mm7107e2

10. Tartof SY, Slezak JM, Fischer H, et al. Effectiveness of mRNA BNT162b2 COVID-19 vaccine up to 6 months in a large integrated health system in the USA: a retrospective cohort study. Lancet. 10 16 2021;398(10309):1407–1416. doi:10.1016/S0140-6736(21)02183-8

11. Jones JM, Opsomer JD, Stone M, et al. Updated US Infection- and Vaccine-Induced SARS-CoV-2 Seroprevalence Estimates Based on Blood Donations, July 2020-December 2021. JAMA. 07 19 2022;328(3):298–301. doi:10.1001/jama.2022.9745

12. Centers for Disease Control and Prevention. Nationwide COVID-19 Infection-Induced Antibody Seroprevalence (Commercial laboratories). Available at: https://covid.cdc.gov/covid-data-tracker/#national-lab. Accessed on: 20 Sept 2022.

13. U.S. Food & Drug Administration. Coronavirus (COVID-19) Update: FDA Authorizes Second Booster Dose of Two COVID-19 Vaccines for Older and Immunocompromised Individuals. Available at: https://www.fda.gov/news-events/press-announcements/coronavirus-covid-19-update-fda-authorizes-second-booster-dose-two-covid-19-vaccines-older-and. Accessed on: 23 June 2022.

14. Centers for Disease Control and Prevention. CDC recommends the first updated COVID-19 booster. Available at: https://www.cdc.gov/media/releases/2022/s0901-covid-19-booster.html. Accessed on: 6 Sept 2022.

15. Thompson MG, Stenehjem E, Grannis S, et al. Effectiveness of Covid-19 Vaccines in Ambulatory and Inpatient Care Settings. N Engl J Med. 10 07 2021;385(15):1355–1371. doi:10.1056/NEJMoa2110362

16. Centers for Disease Control and Prevention. CDC Endorses ACIP’s Updated COVID-19 Vaccine Recommendations. Available at: https://www.cdc.gov/media/releases/2021/s1216-covid-19-vaccines.html. Accessed on: 30 Aug 2022.

17. Embi PJ, Levy ME, Naleway AL, et al. Effectiveness of 2-Dose Vaccination with mRNA COVID-19 Vaccines Against COVID-19-Associated Hospitalizations Among Immunocompromised Adults - Nine States, January-September 2021. MMWR Morb Mortal Wkly Rep. Nov 05 2021;70(44):1553–1559. doi:10.15585/mmwr.mm7044e3

18. Tenforde MW, Self WH, Gaglani M, et al. Effectiveness of mRNA Vaccination in Preventing COVID-19-Associated Invasive Mechanical Ventilation and Death - United States, March 2021-January 2022. MMWR Morb Mortal Wkly Rep. Mar 25 2022;71(12):459–465. doi:10.15585/mmwr.mm7112e1

19. Link-Gelles R, Levy ME, Gaglani M, et al. Effectiveness of 2, 3, and 4 COVID-19 mRNA Vaccine Doses Among Immunocompetent Adults During Periods when SARS-CoV-2 Omicron BA.1 and BA.2/BA.2.12.1 Sublineages Predominated - VISION Network, 10 States, December 2021-June 2022. MMWR Morb Mortal Wkly Rep. Jul 22 2022;71(29):931–939. doi:10.15585/mmwr.mm7129e1

20. Muhsen K, Maimon N, Mizrahi AY, et al. Association of Receipt of the Fourth BNT162b2 Dose With Omicron Infection and COVID-19 Hospitalizations Among Residents of Long-term Care Facilities. JAMA Intern Med. 08 01 2022;182(8):859–867. doi:10.1001/jamainternmed.2022.2658

21. Andrews N, Tessier E, Stowe J, et al. Duration of Protection against Mild and Severe Disease by Covid-19 Vaccines. N Engl J Med. 01 27 2022;386(4):340–350. doi:10.1056/NEJMoa2115481

22. Tenforde MW, Self WH, Adams K, et al. Association Between mRNA Vaccination and COVID-19 Hospitalization and Disease Severity. JAMA. 11 23 2021;326(20):2043–2054. doi:10.1001/jama.2021.19499

23. Centers for Disease Control and Prevention. COVID data tracker. Available at: https://covid.cdc.gov/covid-data-tracker/#datatracker-home. Accessed on: 20 Aug 2021.

24. Feikin DR, Higdon MM, Abu-Raddad LJ, et al. Duration of effectiveness of vaccines against SARS-CoV-2 infection and COVID-19 disease: results of a systematic review and meta-regression. Lancet. Feb 21 2022;doi:10.1016/S0140-6736(22)00152-0

25. Rubin R. COVID-19 Boosters This Fall to Include Omicron Antigen, but Questions Remain About Its Value. JAMA. 08 02 2022;328(5):412–414. doi:10.1001/jama.2022.11252

26. Collie S, Nayager J, Bamford L, Bekker LG, Zylstra M, Gray G. Effectiveness and Durability of the BNT162b2 Vaccine against Omicron Sublineages in South Africa. N Engl J Med. Sep 14 2022;doi:10.1056/NEJMc2210093

