## Supplemental Appendix for "Association between COVID-19 mRNA vaccination and COVID-19 illness and severity during Omicron BA.4 and BA.5 sublineage periods"

**Supplemental Online Content**

**eAppendix 1.** Supplementary Methods.

**eAppendix 2.** Supplementary Tables and Figures.

**eTable 1.** Characteristics of VISION Network Study Sites.

**eTable 2.** COVID-19–Like Illness Categories and Corresponding *International Classification of Diseases, 9th and 10th Revision* Diagnosis Codes.

**eTable 3.** Covariates with Remaining Imbalances Between Vaccinated and Unvaccinated Patients After Application of Inverse Propensity-to-Be-Vaccinated Weighting for Calculation of Adjusted Odds Ratios.

**eTable 4.** Relative Association of COVID-19–Associated Emergency Department or Urgent Care Encounters with Prior Vaccination with Three Versus Two or Four Versus Three mRNA Vaccine Doses, by Age Group.

**eTable 5.** Relative Association of COVID-19–Associated Hospitalization with Prior Vaccination with Three Versus Two or Four Versus Three mRNA Vaccine Doses, by Age Group.

**eFigure 1.** Association of COVID-19–Associated Emergency Department or Urgent Care Encounters with Prior Vaccination with Two, Three, or Four mRNA Vaccine Doses, by mRNA Vaccine Product(s) Received.

**eFigure 2.** Association of COVID-19–Associated Hospitalization with Prior Vaccination with Two, Three, or Four mRNA Vaccine Doses, by mRNA Vaccine Product(s) Received.

**eFigure 3.** Association of COVID-19–Associated Intensive Care Unit Admission and/or In-Hospital Death with Prior Vaccination with Two, Three, or Four mRNA Vaccine Doses, by mRNA Vaccine Product(s) Received.

**eFigure 4.** Association of COVID-19–Associated Emergency Department or Urgent Care Encounters with Prior Vaccination with Two, Three, or Four mRNA Vaccine Doses, by Age Group, Among Patients Without a Prior Documented SARS-CoV-2 Infection.

**eFigure 5.** Association of COVID-19–Associated Hospitalization with Prior Vaccination with Two, Three, or Four mRNA Vaccine Doses, by Age Group, Among Patients Without a Prior Documented SARS-CoV-2 Infection.

**eFigure 6.** Association of COVID-19–Associated Intensive Care Unit Admission and/or In-Hospital Death with Prior Vaccination with Two, Three, or Four mRNA Vaccine Doses, by Age Group, Among Patients Without a Prior Documented SARS-CoV-2 Infection.

**eAppendix 1. Supplementary Methods**

**Section 1: Extracting Percent Positivity Data from HHS Protect**

National laboratory testing data, including data on the state and county level, are available on the password-protected HHS Protect Public Data Hub. The laboratory testing data include viral SARS-CoV-2 laboratory test results (reverse transcription polymerase chain reaction [RT-PCR]) from over 1,000 United States laboratories and testing locations including commercial and reference laboratories, public health laboratories, hospital laboratories, and other testing locations. Data presented in HHS Protect are representative of diagnostic specimens being tested and reflect the majority of, but not all, SARS-CoV-2 laboratory-based testing conducted in the United States. Data from HHS Protect are electronic health records and do not contain personally identifiable information (see <https://www.hhs.gov/sites/default/files/hhs-protect-faqs.pdf> for more information).

For this analysis, daily laboratory testing data were downloaded from HHS Protect and aggregated at the county-level by date of report. Using the county-level average of the daily percentage of tests that were positive during each day and the prior six days, we further aggregated and computed each date’s daily seven-day average separately for each site geographic sub-region (aggregates of counties) by taking into account county population sizes. This population-weighted daily value for each site sub-region was then assigned to each medical encounter as a measure of local SARS-CoV-2 circulation based on the medical encounter index date and site sub-region of the medical facility within which the respective encounter occurred.

**Section 2: Statistical Methods**

**Section 2.1: Overview**

This section further expands on the statistical methodology that was used to estimate the association of symptomatic laboratory-confirmed SARS-CoV-2 infection in an emergency department (ED) or urgent care (UC) clinic setting or hospital setting with vaccination status. In the context of a test-negative case-control study conducted during a period of ≥50% Omicron BA.4/BA.5 sublineage predominance among medical encounters for patients with COVID-19–like illness, the odds of having each specific vaccination status (each defined based on the number of doses received and number of days since the most recent dose) versus unvaccinated status was compared between SARS-CoV-2–positive cases and SARS-CoV-2–negative controls. The test-negative design can minimize biases associated with access to vaccines and healthcare seeking behaviors and has been used extensively to estimate vaccine effectiveness (VE) against medically attended influenza virus illness (1,2). Methods for the current analysis were based on those of prior VISION Network analyses, which have been detailed elsewhere (3). Analyses were conducted separately among ED or UC encounters, hospitalizations, and hospitalizations with intensive care unit (ICU) admission and/or in-hospital death ≤28 days after admission. Analyses were also conducted separately for each pairwise vaccination status comparison (e.g., 3 doses with 3rd dose 7-119 days earlier versus unvaccinated). Both inverse propensity score weighting and covariate adjustment procedures were used to control for confounding, that is, control for differences in characteristics between vaccinated and unvaccinated patients when estimating odds ratios for each pairwise comparison, which were then used to estimate VE in each setting using the formula VE = [1−adjusted OR] x 100%.

**Section 2.2: Repeat Encounters**

Although data were collected and analyzed at the encounter level rather than at the individual patient level, repeat encounters among unique patients were not expected to be highly prevalent given the relatively short time period examined. In general, when the percentage of repeat encounters (for patients who previously contributed an encounter) exceeds 10%, a sensitivity analysis is conducted to assess the effect of within-person correlation on ORs and confidence intervals. In this analysis of ED/UC encounters and hospitalizations during a period of ≥50% Omicron BA.4/BA.5 sublineage predominance, there were 8.5% (n=6,364) repeat ED/UC encounters and 0.4% (n=91) repeat hospitalizations among patients with a prior encounter of the respective type; thus, no further action was taken.

**Section 2.3: Inverse Propensity Score Weighting**

Using established methods for estimating propensity scores within case-control studies (4), we first estimated propensity-for-vaccination scores among SARS-CoV–negative controls, with potential confounding variables used as independent variables and vaccination status as the dependent variable. Next, the fitted model was used to calculate propensity-for-vaccination scores for SARS-CoV-2–positive cases. Because each vaccinated category was compared with unvaccinated status in separate analyses, the propensity score represented the estimated probability of being in the specific vaccination category of interest versus being unvaccinated, conditional on measured covariates representing potential confounding variables. Finally, in primary multivariable regression models to estimate the association between symptomatic medically attended laboratory-confirmed SARS-CoV-2 infection and vaccination status, vaccinated patients were weighted by the inverse of their propensity to be vaccinated and unvaccinated patients were weighted by the inverse of their propensity to not be vaccinated. Inverse propensity score weighting was designed to estimate an overall average treatment effect.

Propensity to be vaccinated was estimated using boosted regression trees (BRT), a nonparametric sequential regression technique (4). Regularization settings to prevent overfitting by BRT methods were determined based on overall sample size; however, the following guidelines were followed: shallow tree depth (2-3 interaction levels), large number of trees (7,000), low learning rate (0.01), and 75% bagging.

Among a set of measured covariates that were identified as potential confounders, those included in the propensity score model were covariates empirically determined to be associated with both the outcome (case-control status) and exposure (vaccination status), with significant differences between groups defined as those with an absolute standardized mean or proportion difference >0.10. The following socio-demographic, facility, and medical factors were considered for inclusion: age, sex, race, ethnicity, Medicaid status, calendar date (number of days since January 1, 2021 based on medical encounter index date), geographic region (based on sub-regions defined for each site), local SARS-CoV-2 circulation on the day of each medical encounter index date, urban-rural classification of facility, hospital type (hospital setting only), number of hospital beds (hospital setting only), chronic respiratory condition, chronic non-respiratory condition, asthma, chronic obstructive pulmonary disease, other chronic lung disease, heart failure, ischemic heart disease, hypertension, other heart disease, stroke, other cerebrovascular disease, diabetes type 1, diabetes type 2, diabetes due to underlying conditions or other specified diabetes, other metabolic disease (excluding diabetes), clinical obesity, clinical underweight, renal disease, liver disease, blood disorder, dementia, other neurological/musculoskeletal disorder, Down syndrome, and the presence of at least one prior molecular or rapid antigen SARS-CoV-2 test record documented in the electronic medical record ≥15 days before the medical encounter index date (pre-vaccination, if vaccinated). Four covariates were included in the propensity score model regardless of their association with the outcome and exposure: age, calendar date, geographic region, and local SARS-CoV-2 circulation on the day of each medical encounter index date.

Applying best practices for inverse probability of treatment weights described by Austin and Stuart (5), the distributions of weights were examined for each vaccination status comparison in each medical setting. In each subgroup, outlying weights were identified at the extreme upper end of the distribution. Therefore, we truncated weights at the 99th percentile for each subgroup. Propensity scores and weights were calculated using the ‘twang’ R package (6). All propensity score analyses were conducted using R version 4.1.2.

**Section 2.4: Primary Outcome Model and Covariate Adjustment**

The primary outcome model to estimate the association between symptomatic medically attended laboratory-confirmed SARS-CoV-2 infection and vaccination status was a multivariable logistic regression model, with SARS-CoV-2 test result (i.e., case-control status) as the dependent variable and vaccination status as a dichotomous independent variable. Medical encounter observations were weighted by their inverse propensity to be vaccinated (if vaccinated) or unvaccinated (if not vaccinated). Four covariates were also directly included as additional independent variables in the regression model to account for possible residual confounding that remained after inverse propensity score weighting based on BRT modeling. The four variables were age (as a spline), calendar date (as spline), geographic region, and local SARS-CoV-2 circulation on the day of each medical encounter index date (as a spline). Spline functions for calendar date, local SARS-CoV-2 circulation, and age were defined as natural cubic splines with knots at quartiles. In addition, any other covariates with distributions that remained imbalanced between vaccinated and unvaccinated patients after inverse propensity score weighting, based on an absolute standardized mean or proportion difference >0.2, were also included directly in the respective regression model. The list of unbalanced variables for each model is presented in eTable 3.

**Section 2.5: Relative VE**

In addition to calculating ORs to estimate absolute VE (i.e., VE for receipt of vaccine compared with unvaccinated status), ORs were also calculated to estimate relative VE, for which a specific vaccinated group was compared with a different vaccinated group in order to determine the incremental benefit of receiving an additional vaccine dose when recommend. Relative VE was estimated by comparing individuals who had recently received a 3^rd^ or 4^th^ dose to those who were eligible for, but had not received, the 3^rd^ or 4^th^ dose, respectively. More specifically, the two comparisons were: (1) 3 doses with the 3^rd^ dose in the last 7-119 days versus 2 doses with the 2^nd^ dose ≥150 days earlier; and, among patients aged ≥50 years, (2) 4 doses with the 4^th^ dose within the last 7-119 days versus 3 doses with the 3^rd^ dose ≥120 days earlier.

To calculate ORs reflecting relative VE comparisons, with receipt of 2 or 3 doses serving as the referent group, a similar methodology was used: patient encounters were weighted based on their inverse propensity to be 3-dose vaccinated (if 3-dose vaccinated) or 2-dose vaccinated (if 2-dose vaccinated) – or 4-dose vaccinated (if 4-dose vaccinated) or 3-dose vaccinated (if 3-dose vaccinated) – and a dichotomous variable for vaccination status (3- versus 2-dose vaccinated or 4- versus 3-dose vaccinated) was included as the independent variable used in primary outcome models.

**Section 2.6: Subgroup Analyses**

The analyses described were conducted in each setting (ED or UC encounters, hospitalizations, and hospitalizations with ICU admission and/or in-hospital death) and in different subgroups within each setting. Analyses were conducted among all adults aged ≥18 years as well as separately among three age groups (18-49, 50-64, and ≥65 years). They were also conducted separately for each vaccine product(s) received (mRNA-1273 [Moderna], BNT162b2 [Pfizer-BioNTech], and heterologous pattern) and among patients without prior infection documented in the electronic medical record ≥15 days prior to the hospital admission or encounter date. Propensity score weights and OR estimates for each vaccination status comparison were only calculated using patient encounters qualifying for inclusion in the respective subgroup.

**References**

1. Foppa IM, Haber M, Ferdinands JM, Shay DK. The case test-negative design for studies of the effectiveness of influenza vaccine. Vaccine. 2013; 31:3104–9.
2. Jackson ML, Nelson JC. The test-negative design for estimating influenza vaccine effectiveness. Vaccine.2013; 31:2165–8.
3. Thompson MG, Stenehjem E, Grannis S, Ball S, et al. Effectiveness of COVID-19 vaccines in ambulatory and inpatient care settings. 2011; 385(15): 1355-71.
4. McCaffrey DF, Ridgeway G, Morral AR. Propensity score estimation with boosted regression for evaluating causal effects in observational studies. Psychological methods. 2004; 9(4): 403.
5. Austin PC, Stuart, EA. Moving towards best practices when using inverse probability of treatment weights (IPTW) using the propensity score to estimate causal treatment effects in observational studies. Statistics and Medicine, 2015; 34: 3661-3679.
6. Ridgeway G, McCaffrey D, Morral AR, Burgette L, Grriffin BA. Toolkit for Weighting and Analysis of Nonequivalent Groups: A tutorial for the twang package. Santa Monica, CA: RAND Corporation; 2017.

**eAppendix 2. Supplementary Tables and Figures**

**eTable 1. Characteristics of VISION Network Study Sites.**

| **Network partner (state)** | **No. geographic sub-regions^a^ (N=45)** | **No. of hospitals (N=268)** | **No. of EDs (N=292)** | **No. of urgent care clinics (N=140)** | **≥50% Omicron BA.1 sublineage predominance period** | **≥50% Omicron BA.2/ BA.2.12.1 sublineage predominance period** | **≥50% Omicron BA.4/BA.5 sublineage predominance period** | **Source of vaccination records** | **Vaccine record lag^b^** |
| --- | --- | --- | --- | --- | --- | --- | --- | --- | --- |
| Baylor Scott & White Health (Texas) | 9^c^ | 26 | 29 | 7 | 12/16/21–3/18/22 | 3/19–6/21/22 | 6/22–8/20/22 | ImmTrac Texas Immunization Registry and EHRs | 3 days |
| Columbia University Irving Medical Center (New York) | 1 | 3 | 2 | N/A | 12/18/21–3/16/22 | 3/17–6/28/22 | 6/29–8/20/22 | New York Citywide Immunization Registry and EHRs | 1 week |
| HealthPartners (Minnesota and Wisconsin) | 2 | 10 | 10 | 23 | 12/25/21–3/21/22 | 3/22–6/21/22 | 6/22–8/20/22 | Minnesota Immunization Information Connection and EHRs | 1 week |
| Intermountain Healthcare (Utah) | 8 | 22 | 22 | 33 | 12/24/21–3/18/22 | 3/19–6/22/22 | 6/23–8/20/22 | Utah State Immunization Information System and EHRs | 1 week |
| Kaiser Permanente Northern California (California) | 8 | 29 | 29 | N/A | 12/21/21–3/20/22 | 3/21–6/24/22 | 6/25–8/20/22 | California Immunization Registry, CARE Everywhere^d^, pharmacy data, claims data, and EHRs | 2 weeks |
| Kaiser Permanente Northwest (Oregon and Washington) | 3 | 57 | 97 | 57 | 12/24/21–3/23/22 | 3/24–6/28/22 | 6/29–8/20/22 | Oregon Immunization Information System, Washington State Immunization Information System, claims data, and EHRs | 2 weeks |
| Paso del Norte Health Information Exchange (Texas) | 1 | 7 | N/A | N/A | 12/29/21–3/29/22 | 3/30–6/21/22 | 6/22–8/20/22 | EHRs | 1 week |
| Regenstrief Institute (Indiana) | 10^e^ | 102 | 103 | N/A | 12/26/21–3/20/22 | 3/21–6/18/22 | 6/19–8/20/22 | Children and Hoosier Immunization Registry Program | 1 week |
| University of Colorado (Colorado) | 3 | 12 | N/A | 20 | 12/19/21–3/20/22 | 3/21–6/18/22 | 6/19–8/20/22 | Colorado Immunization Information System and EHRs | 1 week |

Abbreviations: ED, emergency department; EHRs, electronic health records; N/A, not applicable; UC, urgent care.

^a^ Each site defined sub-regions that represent meaningfully distinct geographic areas within their network. Sub-region values were assigned to medical encounters based on the location of the admitting hospital, ED, or UC clinic and were used for purposes of adjustment for geographic region in multivariable regression modeling.

^b^ The vaccine record lag is the duration of time post-vaccination before vaccine records are expected to be available in sites’ records contributing to data collection for this study.

^c^ For Baylor Scott & White Health, inpatient facilities were located in only 8 of the 9 sub-regions.

^d^ CARE Everywhere is an Epic electronic health record inter-hospital system for vaccination record sharing.

^e^ For Regenstrief Institute, one of the 10 sub-region values represents unknown location of facility, and inpatient facilities were located in only 8 of the 9 defined sub-regions.

**eTable 2. COVID-19–Like Illness Categories and Corresponding *International Classification of Diseases, 9th and 10th Revision* Diagnosis Codes.**

| **Description of Diagnosis** | **ICD-10 codes** | **ICD-9 codes** |
| --- | --- | --- |
| **COVID-19 Pneumonia** |  |  |
| Pneumonia due to SARS-associated coronavirus | J12.81 | N/A |
| Pneumonia due to coronavirus disease 2019 | J12.82 | N/A |
| **Influenza Pneumonia** |  |  |
| Influenza due to identified novel influenza A virus with pneumonia | J09.X1 | 488.81 |
| Influenza due to other identified influenza virus with pneumonia | J10.0* | N/A |
| Influenza due to other identified influenza virus with unspecified type of pneumonia | J10.00 | 487.0 |
| Influenza due to other identified influenza virus with the same other identified influenza virus pneumonia | J10.01 | 487.0 |
| Influenza due to other identified influenza virus with other specified pneumonia | J10.08 | 487.0, 488.11 |
| Influenza due to unidentified influenza virus with pneumonia | J11.0* | N/A |
| Influenza due to unidentified influenza virus with unspecified type of pneumonia | J11.00 | 487.0 |
| Influenza due to unidentified influenza virus with specified pneumonia | J11.08 | 487.0 |
| Influenza with pneumonia | N/A | 487* |
| **Other Viral Pneumonia** | J12.0, J12.1, J12.3, J12.3, J12.89, J12.9 | 480* |
| **Bacterial and Other Pneumonia** |  |  |
| Streptococcus pneumoniae pneumonia | J13 | 481 |
| Hemophilus influenzae pneumonia | J14 | 482.2 |
| Other bacterial pneumonia | J15* | 482* |
| Pneumonia due to other specified organism | J16* | 483* |
| Pneumonia in infectious diseases classified elsewhere | J17 | 484* |
| Pneumonia, unspecified organism | J18* | 486 |
| **Influenza Disease** | J09.X2, J09.X3, J09.X9, J10.1, J10.2, J10.8*, J11.1, J11.2, J11.8* | 488* |
| **Acute respiratory distress syndrome** | J80 | 518.82 |
| **COPD with acute exacerbation** | J44.1 | 491.21 |
| **Asthma acute exacerbation** | J45.21, J45.22, J45.31, J45.32, J45.41, J45.42, J45.51, J45.52, J45.901, J45.902 | 493.01, 493.02, 493.11, 493.12, 493.21, 493.22, 493.91, 493.92 |
| **Respiratory failure** |  |  |
| Acute respiratory failure | J96.0* | 518.81 |
| Acute and chronic respiratory failure | J96.2* | 518.84 |
| Respiratory arrest | R09.2 | 799.1 |
| **Other acute lower respiratory tract infections** |  |  |
| Acute bronchitis | J20* | 466.0 |
| Acute bronchiolitis | J21* | 466.1* |
| Unspecified acute lower respiratory infection | J22 | 519.8 |
| Bronchitis, not specified as acute or chronic | J40 | 490 |
| COPD with acute lower respiratory infection | J44.0 | 491.22 |
| Simple and mucopurulent chronic bronchitis | J41* | 491* |
| Unspecified chronic bronchitis | J42 | 491.9 |
| Emphysema | J43* | 492* |
| Bronchiectasis | J47* | 494* |
| Abscess of lung and mediastinum | J85* | 513* |
| Gangrene and necrosis of lung | J85.0 | N/A |
| Abscess of lung without pneumonia | J85.2 | 513.0 |
| Abscess of mediastinum | J85.3 | 513.1 |
| Abscess of lung with pneumonia | J85.1 | 513.0 |
| Pyothorax | J86* | 510* |
| **Acute and chronic sinusitis** | J01* , J32* | 461* , 473* |
| **Acute upper respiratory tract infections** | J00*, J02*, J03*, J04*, J05*, J06* | 460*, 462, 463, 464*, 465* |
| **Signs and symptoms of acute respiratory illness** |  |  |
| Hemoptysis | R04.2 | 786.3 |
| Cough | R05 R05.1, R05.2, R05.4, R05.8, R05.9 | 786.2 |
| Dyspnea unspecified | R06.00 | 786.09 |
| Shortness of breath | R06.02 | 786.05 |
| Acute respiratory distress | R06.03 | N/A |
| Stridor | R06.1 | 786.1 |
| Wheezing | R06.2 | 786.07 |
| Other abnormalities of breathing | R06.8 | N/A |
| Apnea, not elsewhere classified | R06.81 | 786.03 |
| Tachypnea, not elsewhere classified | R06.82 | 786.06 |
| Other abnormalities of breathing/ Other symptoms involving head & neck | R06.89 | 784.99 |
| Other dyspnea and respiratory abnormality | N/A | 786.09 |
| Other symptoms involving respiratory system and chest | N/A | 786.9 |
| Chest pain on breathing/ painful respiration | R07.1 | 786.52 |
| Asphyxia and hypoxemia | R09.0* | N/A |
| Asphyxia | R09.01 | 799.01 |
| Hypoxemia | R09.02 | 799.02 |
| Pleurisy | R09.1 | 511.0 |
| Respiratory arrest | R09.2 | 799.1 |
| Abnormal sputum | R09.3 | 786.4 |
| Other specified symptoms and signs involving the circulatory and respiratory systems | R09.8* | 478.19, 784.91, 786.7 |
| **Signs and symptoms of acute febrile illness** |  |  |
| Fever | R50* | N/A |
| Fever presenting with conditions classified elsewhere | R50.81 | 780.61 |
| Fever unspecified | R50.9 | 780.6 |
| Chills (without fever) | R68.83 | 780.64 |
| **Signs and symptoms of acute non-respiratory illness** |  |  |
| Diarrhea | R19.7 | 787.91 |
| Disturbance of smell and taste | R43* | N/A |
| Unspecified disturbances of smell and taste | R43.9 | 781.1, V41.5 |
| Headache | R51.9 | 784.0 |
| Myalgia | M79.10, M79.18 | 729.1 |
| Sepsis - Symptoms and signs specifically associated with systemic inflammation and infection | R65* | 785.52 |
| Other malaise | R53.81 | 780.79 |
| Other fatigue | R53.83 | 780.79 |
| Shock, unspecified | R57.9 | 785.5 |
| Debility unspecified | N/A | 799.3 |
| Altered level of consciousness / altered mental status | R41.82, R40.0, R40.1 | 780.97, 780.0* |
| Weakness | R53.1 | 780.79 |
| Nausea and Vomiting | R11.0, R11.10, R11.11, R11.15, R11.2 | 787* |
| Rash and other nonspecific skin eruption | R21* | 782.1 |
| Abdominal pain | R10.0, R10.1*, R10.2, R10.3*, R10.81*, R10.84, R10.9 | 789* |

Abbreviations: COPD, Chronic Obstructive Pulmonary Disease; ICD-10, *International Classification of Diseases, 10^th^ Revision*; ICD-9, *International Classification of Diseases, 9^th^ Revision*; N/A, not applicable.

*Includes all sub-codes.

**eTable 3. Covariates with Remaining Imbalances Between Vaccinated and Unvaccinated Patients After Application of Inverse Propensity-to-Be-Vaccinated Weighting for Calculation of Adjusted Odds Ratios.**

| **Vaccination status (each compared with unvaccinated)** | **Setting** | **Age group, y** | **Covariates with absolute SMD >0.2 after weighting^a^** |
| --- | --- | --- | --- |
| 2 doses (14-149 days earlier) | ED or UC encounters | ≥18 | None |
| 2 doses (≥150 days earlier) | ED or UC encounters | ≥18 | None |
| 3 doses (7-119 days earlier) | ED or UC encounters | ≥18 | None |
| 3 doses (≥120 days earlier) | ED or UC encounters | ≥18 | None |
| 2 doses (14-149 days earlier) | ED or UC encounters | 18-49 | None |
| 2 doses (≥150 days earlier) | ED or UC encounters | 18-49 | None |
| 3 doses (7-119 days earlier) | ED or UC encounters | 18-49 | None |
| 3 doses (≥120 days earlier) | ED or UC encounters | 18-49 | None |
| 2 doses (14-149 days earlier) | ED or UC encounters | 50-64 | None |
| 2 doses (≥150 days earlier) | ED or UC encounters | 50-64 | None |
| 3 doses (7-119 days earlier) | ED or UC encounters | 50-64 | None |
| 3 doses (≥120 days earlier) | ED or UC encounters | 50-64 | None |
| 4 doses (7-59 days earlier) | ED or UC encounters | 50-64 | Hypertension (SMD=0.21) |
| 4 doses (≥60 days earlier) | ED or UC encounters | 50-64 | Presence of prior SARS-CoV-2 test record (SMD=0.25) |
| 2 doses (14-149 days earlier) | ED or UC encounters | ≥65 | None |
| 2 doses (≥150 days earlier) | ED or UC encounters | ≥65 | None |
| 3 doses (7-119 days earlier) | ED or UC encounters | ≥65 | None |
| 3 doses (≥120 days earlier) | ED or UC encounters | ≥65 | None |
| 4 doses (7-59 days earlier) | ED or UC encounters | ≥65 | None |
| 4 doses (≥60 days earlier) | ED or UC encounters | ≥65 | None |
| 2 doses (14-149 days earlier) | Hospitalizations | ≥18 | None |
| 2 doses (≥150 days earlier) | Hospitalizations | ≥18 | None |
| 3 doses (7-119 days earlier) | Hospitalizations | ≥18 | None |
| 3 doses (≥120 days earlier) | Hospitalizations | ≥18 | None |
| 2 doses (14-149 days earlier) | Hospitalizations | 18-49 | Medicaid status (SMD=0.25) |
| 2 doses (≥150 days earlier) | Hospitalizations | 18-49 | None |
| 3 doses (7-119 days earlier) | Hospitalizations | 18-49 | Chronic non-respiratory condition (SMD=0.40), hypertension (SMD=0.24), other metabolic disease (SMD=0.24) |
| 3 doses (≥120 days earlier) | Hospitalizations | 18-49 | None |
| 2 doses (14-149 days earlier) | Hospitalizations | 50-64 | Medicaid status (SMD=0.24) |
| 2 doses (≥150 days earlier) | Hospitalizations | 50-64 | None |
| 3 doses (7-119 days earlier) | Hospitalizations | 50-64 | None |
| 3 doses (≥120 days earlier) | Hospitalizations | 50-64 | None |
| 4 doses (7-59 days earlier) | Hospitalizations | 50-64 | Medicaid status (SMD=0.34), heart failure (SMD=0.28) |
| 4 doses (≥60 days earlier) | Hospitalizations | 50-64 | Race (SMD=0.29), Medicaid status (SMD=0.24), other chronic lung disease (SMD=0.27), heart failure (SMD=0.26) |
| 2 doses (14-149 days earlier) | Hospitalizations | ≥65 | Chronic obstructive pulmonary disease (SMD=0.33) |
| 2 doses (≥150 days earlier) | Hospitalizations | ≥65 | None |
| 3 doses (7-119 days earlier) | Hospitalizations | ≥65 | Presence of prior SARS-CoV-2 test record (SMD=0.30) |
| 3 doses (≥120 days earlier) | Hospitalizations | ≥65 | None |
| 4 doses (7-59 days earlier) | Hospitalizations | ≥65 | None |
| 4 doses (≥60 days earlier) | Hospitalizations | ≥65 | None |
| 2 doses (14-149 days earlier) | ICU admission or in-hospital death | ≥18 | None |
| 2 doses (≥150 days earlier) | ICU admission or in-hospital death | ≥18 | None |
| 3 doses (7-119 days earlier) | ICU admission or in-hospital death | ≥18 | None |
| 3 doses (≥120 days earlier) | ICU admission or in-hospital death | ≥18 | None |
| 2 doses (14-149 days earlier) | ICU admission or in-hospital death | 18-49 | Medicaid status (SMD=0.25) |
| 2 doses (≥150 days earlier) | ICU admission or in-hospital death | 18-49 | None |
| 3 doses (7-119 days earlier) | ICU admission or in-hospital death | 18-49 | Medicaid status (SMD=0.20), chronic non-respiratory condition (SMD=0.35), hypertension (SMD=0.24), other heart disease (SMD=0.26) |
| 3 doses (≥120 days earlier) | ICU admission or in-hospital death | 18-49 | None |
| 2 doses (14-149 days earlier) | ICU admission or in-hospital death | 50-64 | Chronic respiratory condition (SMD=0.21), presence of prior SARS-CoV-2 test record (SMD=0.35) |
| 2 doses (≥150 days earlier) | ICU admission or in-hospital death | 50-64 | None |
| 3 doses (7-119 days earlier) | ICU admission or in-hospital death | 50-64 | Sex (SMD=0.21), hospital type (SMD=0.34) |
| 3 doses (≥120 days earlier) | ICU admission or in-hospital death | 50-64 | None |
| 4 doses (7-59 days earlier) | ICU admission or in-hospital death | 50-64 | Medicaid status (SMD=0.36), heart failure (SMD=0.29), ischemic heart disease (SMD=0.21), presence of prior SARS-CoV-2 test record (SMD=0.37) |
| 4 doses (≥60 days earlier) | ICU admission or in-hospital death | 50-64 | Race (SMD=0.29), Medicaid status (SMD=0.24), other chronic lung disease (SMD=0.28), heart failure (SMD=0.26), clinical underweight (SMD=0.21), other neurological/musculoskeletal disorder (SMD=0.34), presence of prior SARS-CoV-2 test record (SMD=0.36) |
| 2 doses (14-149 days earlier) | ICU admission or in-hospital death | ≥65 | Sex (SMD=0.26), chronic obstructive pulmonary disease (SMD=0.21), other neurological/musculoskeletal disorder (SMD=0.36) |
| 2 doses (≥150 days earlier) | ICU admission or in-hospital death | ≥65 | None |
| 3 doses (7-119 days earlier) | ICU admission or in-hospital death | ≥65 | Presence of prior SARS-CoV-2 test record (SMD=0.31) |
| 3 doses (≥120 days earlier) | ICU admission or in-hospital death | ≥65 | None |
| 4 doses (7-59 days earlier) | ICU admission or in-hospital death | ≥65 | None |
| 4 doses (≥60 days earlier) | ICU admission or in-hospital death | ≥65 | None |

Abbreviations: ED, emergency department; ICU, intensive care unit; SMD, standardized mean or proportion difference; UC, urgent care.

^a^ Covariates included as independent variables in all primary outcome multivariable regression models were age (as a spline), calendar date (as spline), geographic region, and local SARS-CoV-2 circulation on the day of each medical encounter index date (as a spline). Additional covariates evaluated for imbalances after inverse propensity-to-be-vaccinated weighting included sex, race, ethnicity, Medicaid status, urban-rural classification of facility, hospital type (if relevant), number of hospital beds (if relevant), chronic respiratory condition, chronic non-respiratory condition, asthma, chronic obstructive pulmonary disease, other chronic lung disease, heart failure, ischemic heart disease, hypertension, other heart disease, stroke, other cerebrovascular disease, diabetes type 1, diabetes type 2, diabetes due to underlying conditions or other specified diabetes, other metabolic disease (excluding diabetes), clinical obesity, clinical underweight, renal disease, liver disease, blood disorder, dementia, other neurological/musculoskeletal disorder, Down syndrome, and the presence of at least one prior molecular or rapid antigen SARS-CoV-2 test record documented in the electronic medical record ≥15 days before the medical encounter date (pre-vaccination, if vaccinated). An absolute SMD >0.20 indicated a non-negligible difference in variable distributions between vaccinated and unvaccinated patients. All covariates with an absolute SMD >0.20 after weighting were also included in primary regression models for the respective odds ratio estimate(s) to minimize residual confounding.

**eTable 4. Relative Association of COVID-19–Associated Emergency Department or Urgent Care Encounters with Prior Vaccination with Three Versus Two or Four Versus Three mRNA Vaccine Doses, by Age Group.**

| **Encounter type/comparison type/age group/specific vaccination status comparison** | **Total** | **SARS-CoV-2–positive, No. (%)** | **Days since recent dose, median (IQR)** | **Unadjusted OR (95% CI)** | **Adjusted OR^a^ (95% CI)** | **Adjusted VE^b^ % (95% CI)** |
| --- | --- | --- | --- | --- | --- | --- |
| **ED or UC encounters** |  |  |  |  |  |  |
| **Comparisons to assess incremental benefit of additional dose when recommended** |  |  |  |  |  |  |
| **All adults** |  |  |  |  |  |  |
| **3 doses vs. 2 doses** |  |  |  |  |  |  |
| 2 doses ≥150 days earlier (Ref) | 19,594 | 4,436 (22.6) | 424 (326 - 470) | — | — | — |
| 3 doses 7–119 days earlier | 1,539 | 175 (11.4) | 77 (46 - 100) | 0.44 (0.37 - 0.51) | 0.51 (0.42 - 0.61) | 49 (39 - 58) |
| **18-49 years** |  |  |  |  |  |  |
| **3 doses vs. 2 doses** |  |  |  |  |  |  |
| 2 doses ≥150 days earlier (Ref) | 10,409 | 2,217 (21.3) | 399 (305 - 446) | — | — | — |
| 3 doses 7–119 days earlier | 619 | 66 (10.7) | 74 (43 - 101) | 0.44 (0.34 - 0.57) | 0.52 (0.40 - 0.68) | 48 (32 - 60) |
| **50-64 years** |  |  |  |  |  |  |
| **3 doses vs. 2 doses** |  |  |  |  |  |  |
| 2 doses ≥150 days earlier (Ref) | 4,080 | 985 (24.1) | 428 (336 - 466) | — | — | — |
| 3 doses 7–119 days earlier | 360 | 41 (11.4) | 79 (51 - 102) | 0.40 (0.29 - 0.56) | 0.52 (0.37 - 0.73) | 48 (27 - 63) |
| **4 doses vs. 3 doses** |  |  |  |  |  |  |
| 3 doses ≥120 days earlier (Ref) | 4,978 | 1,069 (21.5) | 220 (192 - 248) | — | — | — |
| 4 doses 7–119 days earlier | 1,452 | 210 (14.5) | 66 (41 - 89) | 0.62 (0.53 - 0.73) | 0.77 (0.65 - 0.92) | 23 (8 - 35) |
| **≥65 years** |  |  |  |  |  |  |
| **3 doses vs. 2 doses** |  |  |  |  |  |  |
| 2 doses ≥150 days earlier (Ref) | 5,105 | 1,234 (24.2) | 469 (417 - 499) | — | — | — |
| 3 doses 7–119 days earlier | 560 | 68 (12.1) | 77 (48 - 99) | 0.43 (0.33 - 0.56) | 0.56 (0.41 - 0.77) | 44 (23 - 59) |
| **4 doses vs. 3 doses** |  |  |  |  |  |  |
| 3 doses ≥120 days earlier (Ref) | 9,881 | 2,134 (21.6) | 242 (212 - 268) | — | — | — |
| 4 doses 7–119 days earlier | 5,728 | 821 (14.3) | 75 (50 - 93) | 0.61 (0.56 - 0.66) | 0.65 (0.59 - 0.72) | 35 (28 - 41) |

Abbreviations: CI, confidence interval; ED, emergency department; IQR, interquartile range; OR, odds ratio; Ref, referent group; UC, urgent care; VE, vaccine effectiveness.

^a^ An adjusted OR <1.0 indicates that COVID-19–associated ED/UC encounters were associated with being 2-dose versus 3-dose vaccinated or being 3-dose versus 4-dose vaccinated. ORs were adjusted for age, geographic region, calendar time (days since January 1, 2021), and local virus circulation (percentage of SARS-CoV-2–positive results from testing within the counties surrounding the facility on the date of the encounter) and weighted for inverse propensity to be 3-dose or 2-dose vaccinated or 4-dose or 3-dose vaccinated (calculated separately for each OR estimate). Generalized boosted regression trees were used to estimate the propensity to be 3-dose or 4-dose vaccinated based on the following socio-demographic, facility, and medical factors: age, sex, race, ethnicity, Medicaid status, calendar date, geographic region, local SARS-CoV-2 circulation on the day of each medical visit, urban-rural classification of facility, chronic respiratory condition, chronic non-respiratory condition, asthma, chronic obstructive pulmonary disease, other chronic lung disease, heart failure, ischemic heart disease, hypertension, other heart disease, stroke, other cerebrovascular disease, diabetes type 1, diabetes type 2, diabetes due to underlying conditions or other specified diabetes, other metabolic disease (excluding diabetes), clinical obesity, clinical underweight, renal disease, liver disease, blood disorder, dementia, other neurological/musculoskeletal disorder, Down syndrome, and the presence of at least one prior molecular or rapid antigen SARS-CoV-2 test record documented in the electronic medical record ≥15 days before the medical encounter date (pre-vaccination).

^b^ Relative vaccine effectiveness for prevention of COVID-19–associated ED/UC encounters can be estimated from the adjusted ORs presented in this table with the equation: vaccine effectiveness = (1-adjusted OR) x 100%.

**eTable 5. Relative Association of COVID-19–Associated Hospitalization with Prior Vaccination with Three Versus Two or Four Versus Three mRNA Vaccine Doses, by Age Group.**

| **Encounter type/comparison type/age group/specific vaccination status comparison** | **Total** | **SARS-CoV-2–positive, No. (%)** | **Days since recent dose, median (IQR)** | **Unadjusted OR (95% CI)** | **Adjusted OR^a^ (95% CI)** | **Adjusted VE^b^ % (95% CI)** |
| --- | --- | --- | --- | --- | --- | --- |
| **Hospitalizations** |  |  |  |  |  |  |
| **Comparisons to assess incremental benefit of additional dose when recommended** |  |  |  |  |  |  |
| **All adults** |  |  |  |  |  |  |
| **3 doses vs. 2 doses** |  |  |  |  |  |  |
| 2 doses ≥150 days earlier (Ref) | 4,845 | 824 (17.0) | 450 (373 - 491) | — | — | — |
| 3 doses 7–119 days earlier | 429 | 33 (7.7) | 76 (43 - 100) | 0.41 (0.28 - 0.58) | 0.43 (0.28 - 0.65) | 57 (35 - 72) |
| **18-49 years** |  |  |  |  |  |  |
| **3 doses vs. 2 doses^c^** |  |  |  |  |  |  |
| 2 doses ≥150 days earlier (Ref) | 887 | 109 (12.3) | 399 (304 - 448) | — | — | — |
| 3 doses 7–119 days earlier^d^ | — | — | — | — | — | — |
| **50-64 years** |  |  |  |  |  |  |
| **3 doses vs. 2 doses^c^** |  |  |  |  |  |  |
| 2 doses ≥150 days earlier (Ref) | 1,130 | 159 (14.1) | 427 (336 - 465) | — | — | — |
| 3 doses 7–119 days earlier^d^ | — | — | — | — | — | — |
| **4 doses vs. 3 doses^c^** |  |  |  |  |  |  |
| 3 doses ≥120 days earlier (Ref) | 1,121 | 133 (11.9) | 220 (194 - 248) | — | — | — |
| 4 doses 7–119 days earlier | 273 | 25 (9.2) | 70 (43 - 89) | 0.75 (0.48 - 1.17) | — | — |
| **≥65 years** |  |  |  |  |  |  |
| **3 doses vs. 2 doses** |  |  |  |  |  |  |
| 2 doses ≥150 days earlier (Ref) | 2,828 | 556 (19.7) | 473 (422 - 503) | — | — | — |
| 3 doses 7–119 days earlier | 289 | 26 (9.0) | 72 (42 - 98) | 0.40 (0.27 - 0.61) | 0.41 (0.25 - 0.67) | 59 (33 - 75) |
| **4 doses vs. 3 doses** |  |  |  |  |  |  |
| 3 doses ≥120 days earlier (Ref) | 4,838 | 913 (18.9) | 240 (211 - 266) | — | — | — |
| 4 doses 7–119 days earlier | 2,179 | 277 (12.7) | 73 (48 - 91) | 0.63 (0.54 - 0.72) | 0.63 (0.54 - 0.75) | 37 (25 - 46) |

Abbreviations: CI, confidence interval; IQR,, interquartile range; OR, odds ratio; Ref, referent group; VE, vaccine effectiveness.

^a^ An adjusted OR <1.0 indicates that COVID-19–associated hospitalization was associated with being 2-dose versus 3-dose vaccinated or being 3-dose versus 4-dose vaccinated. ORs were adjusted for age, geographic region, calendar time (days since January 1, 2021), and local virus circulation (percentage of SARS-CoV-2–positive results from testing within the counties surrounding the facility on the date of the encounter) and weighted for inverse propensity to be 3-dose or 2-dose vaccinated or 4-dose or 3-dose vaccinated (calculated separately for each OR estimate). Generalized boosted regression trees were used to estimate the propensity to be 3-dose or 4-dose vaccinated based on the following socio-demographic, facility, and medical factors: age, sex, race, ethnicity, Medicaid status, calendar date, geographic region, local SARS-CoV-2 circulation on the day of each medical visit, urban-rural classification of facility, hospital type, number of hospital beds, chronic respiratory condition, chronic non-respiratory condition, asthma, chronic obstructive pulmonary disease, other chronic lung disease, heart failure, ischemic heart disease, hypertension, other heart disease, stroke, other cerebrovascular disease, diabetes type 1, diabetes type 2, diabetes due to underlying conditions or other specified diabetes, other metabolic disease (excluding diabetes), clinical obesity, clinical underweight, renal disease, liver disease, blood disorder, dementia, other neurological/musculoskeletal disorder, Down syndrome, and the presence of at least one prior molecular or rapid antigen SARS-CoV-2 test record documented in the electronic medical record ≥15 days before the medical encounter date (pre-vaccination).

^b^ Relative vaccine effectiveness for prevention of COVID-19–associated hospitalization can be estimated from the adjusted ORs presented in this table with the equation: vaccine effectiveness = (1-adjusted OR) x 100%.

^c^ Adjusted ORs and VE estimates are not shown for vaccination status comparisons with confidence intervals greater than 50 percentage points around the VE estimate.

^d^ In vaccination status subgroups with <10 SARS-CoV-2–positive cases, all numbers in the row were removed because of small cell sizes.

**eFigure 1. Association of COVID-19–Associated Emergency Department or Urgent Care Encounters with Prior Vaccination with Two, Three, or Four mRNA Vaccine Doses, by mRNA Vaccine Product(s) Received.**

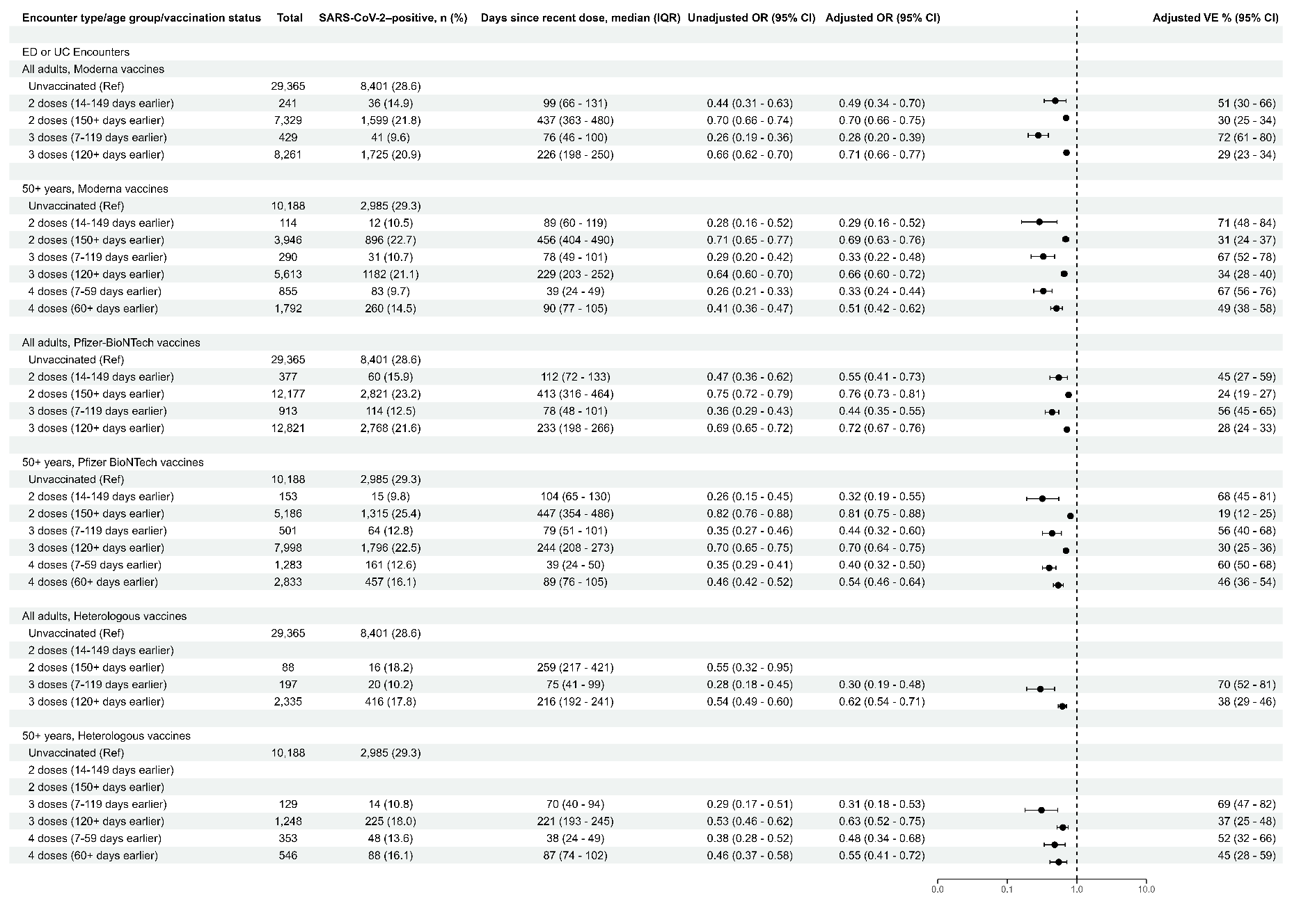

An adjusted OR <1.0 indicates that COVID-19–associated ED/UC encounter was associated with being unvaccinated compared with being vaccinated. ORs were adjusted for age, geographic region, calendar time (days since January 1, 2021), and local virus circulation (percentage of SARS-CoV-2–positive results from testing within the counties surrounding the facility on the date of the encounter) and weighted for inverse propensity to be vaccinated or unvaccinated (calculated separately for each OR estimate). Generalized boosted regression trees were used to estimate the propensity to be vaccinated based on the following socio-demographic, facility, and medical factors: age, sex, race, ethnicity, Medicaid status, calendar date, geographic region, local SARS-CoV-2 circulation on the day of each medical visit, urban-rural classification of facility, chronic respiratory condition, chronic non-respiratory condition, asthma, chronic obstructive pulmonary disease, other chronic lung disease, heart failure, ischemic heart disease, hypertension, other heart disease, stroke, other cerebrovascular disease, diabetes type 1, diabetes type 2, diabetes due to underlying conditions or other specified diabetes, other metabolic disease (excluding diabetes), clinical obesity, clinical underweight, renal disease, liver disease, blood disorder, dementia, other neurological/musculoskeletal disorder, Down syndrome, and the presence of at least one prior molecular or rapid antigen SARS-CoV-2 test record documented in the electronic medical record ≥15 days before the medical encounter date (pre-vaccination, if vaccinated). Vaccine effectiveness for prevention of COVID-19–associated ED/UC encounter can be estimated from the adjusted ORs presented in this table with the equation: vaccine effectiveness = (1-adjusted OR) x 100%. Adjusted ORs and VE estimates are not shown for vaccination status comparisons with confidence intervals greater than 50 percentage points around the VE estimate. Adjusted ORs and VE estimates could not be calculated for the following subgroups due to lack of model convergence: all adults, heterologous vaccines, 2 doses (14-149 days earlier); and 50+ years, heterologous vaccines, 2 doses (14-149 days earlier). In vaccination status subgroups with <10 SARS-CoV-2–positive cases, all numbers in the row were removed because of small cell sizes. CI indicates confidence interval; ED, emergency department; IQR, interquartile range; OR, odds ratio; Ref, referent group; UC, urgent care; VE, vaccine effectiveness.

**eFigure 2. Association of COVID-19–Associated Hospitalization with Prior Vaccination with Two, Three, or Four mRNA Vaccine Doses, by mRNA Vaccine Product(s) Received.**

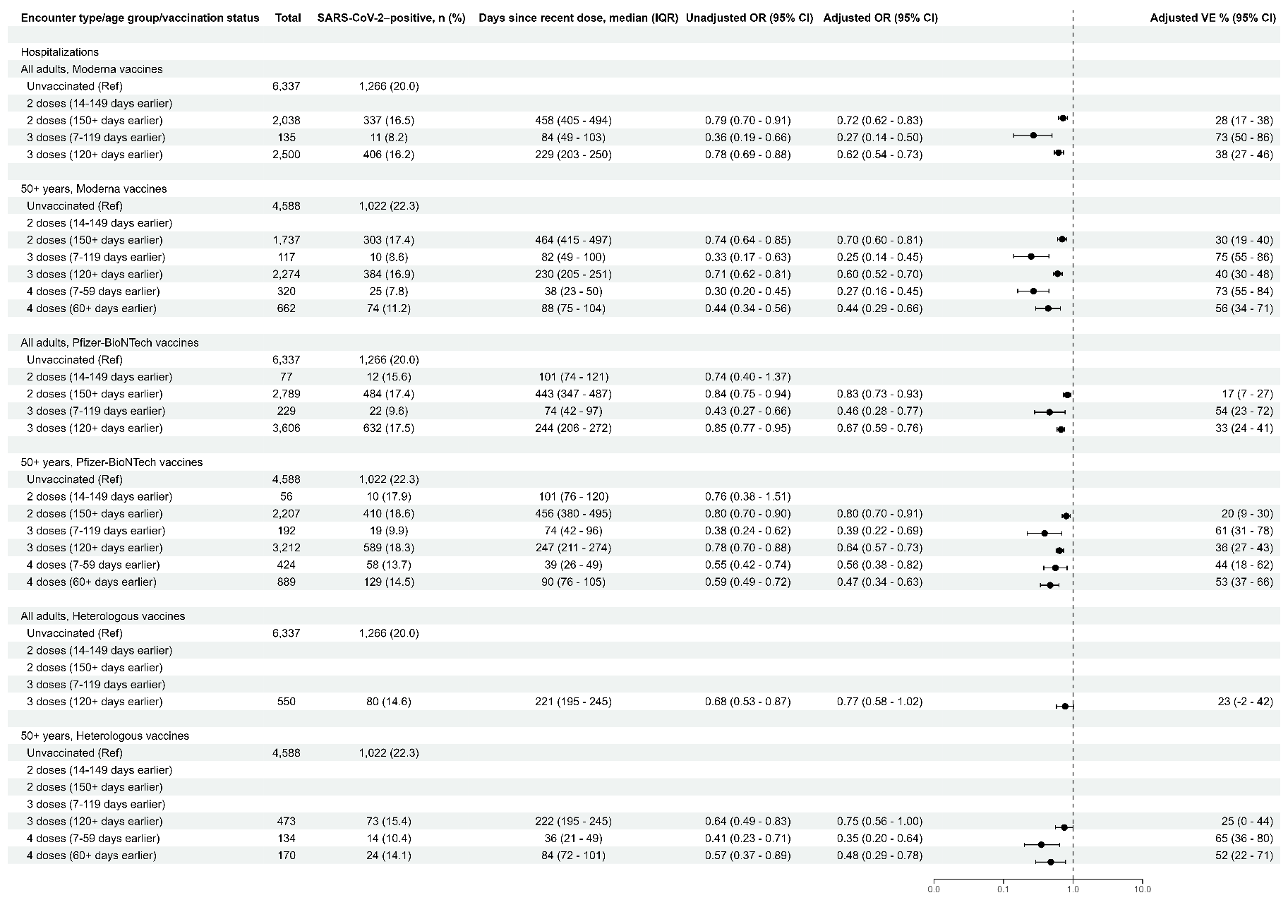

An adjusted OR <1.0 indicates that COVID-19–associated hospitalization was associated with being unvaccinated compared with being vaccinated. ORs were adjusted for age, geographic region, calendar time (days since January 1, 2021), and local virus circulation (percentage of SARS-CoV-2–positive results from testing within the counties surrounding the facility on the date of the encounter) and weighted for inverse propensity to be vaccinated or unvaccinated (calculated separately for each OR estimate). Generalized boosted regression trees were used to estimate the propensity to be vaccinated based on the following socio-demographic, facility, and medical factors: age, sex, race, ethnicity, Medicaid status, calendar date, geographic region, local SARS-CoV-2 circulation on the day of each medical visit, urban-rural classification of facility, hospital type, number of hospital beds, chronic respiratory condition, chronic non-respiratory condition, asthma, chronic obstructive pulmonary disease, other chronic lung disease, heart failure, ischemic heart disease, hypertension, other heart disease, stroke, other cerebrovascular disease, diabetes type 1, diabetes type 2, diabetes due to underlying conditions or other specified diabetes, other metabolic disease (excluding diabetes), clinical obesity, clinical underweight, renal disease, liver disease, blood disorder, dementia, other neurological/musculoskeletal disorder, Down syndrome, and the presence of at least one prior molecular or rapid antigen SARS-CoV-2 test record documented in the electronic medical record ≥15 days before the medical encounter date (pre-vaccination, if vaccinated). Vaccine effectiveness for prevention of COVID-19–associated hospitalization can be estimated from the adjusted ORs presented in this table with the equation: vaccine effectiveness = (1-adjusted OR) x 100%. Adjusted ORs and VE estimates are not shown for vaccination status comparisons with confidence intervals greater than 50 percentage points around the VE estimate. Adjusted ORs and VE estimates could not be calculated for the following subgroups due to lack of model convergence: all adults, heterologous vaccines, 2 doses (14-149 days earlier); all adults, heterologous vaccines, 3 doses (7-119 days earlier); 50+ years, heterologous vaccines, 2 doses (14-149 days earlier); and 50+ years, heterologous vaccines, 3 doses (7-119 days earlier). In vaccination status subgroups with <10 SARS-CoV-2–positive cases, all numbers in the row were removed because of small cell sizes. CI indicates confidence interval; IQR, interquartile range; OR, odds ratio; Ref, referent group; VE, vaccine effectiveness.

**eFigure 3. Association of COVID-19–Associated Intensive Care Unit Admission and/or In-Hospital Death with Prior Vaccination with Two, Three, or Four mRNA Vaccine Doses, by mRNA Vaccine Product(s) Received.**

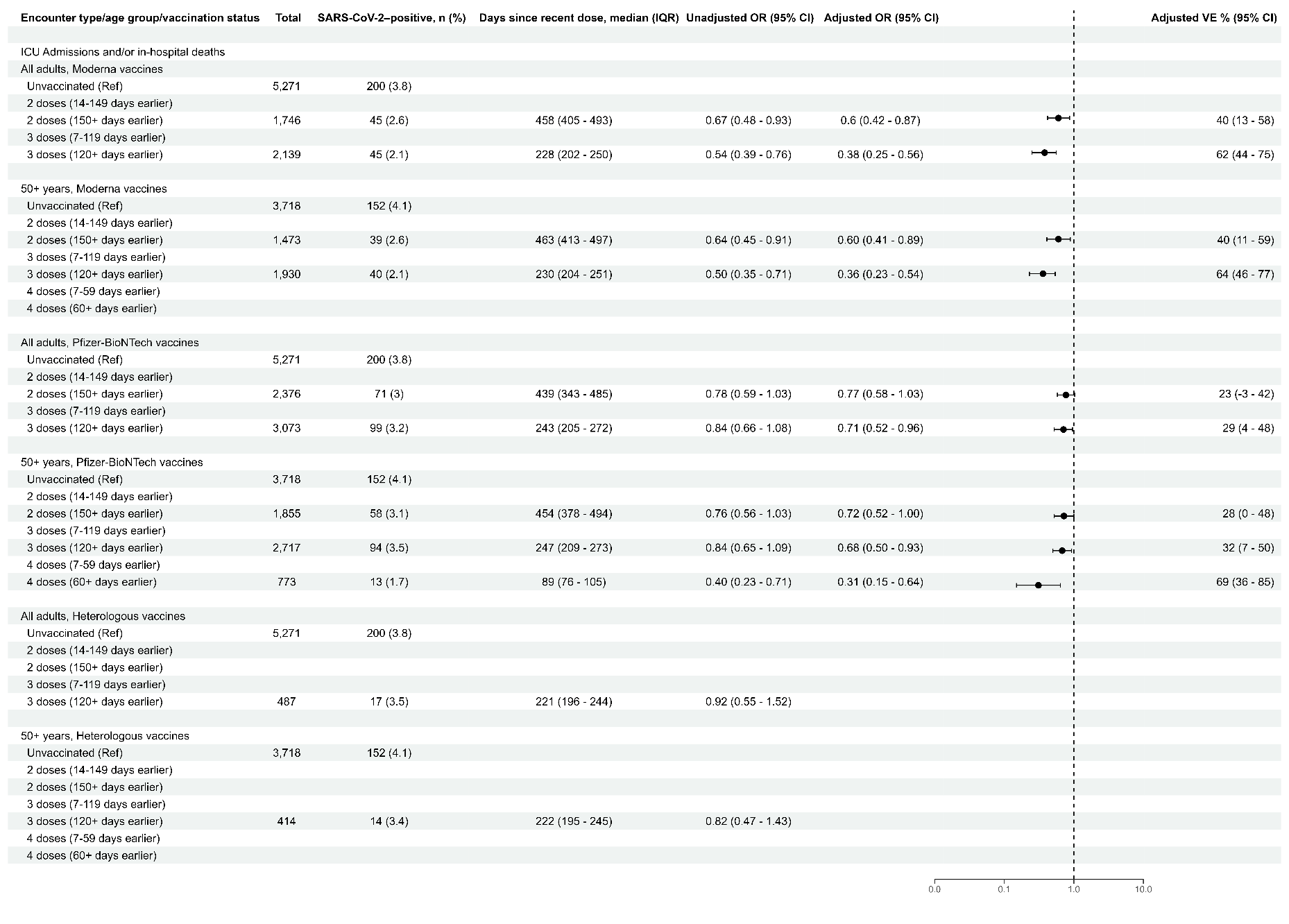

An adjusted OR <1.0 indicates that COVID-19–associated ICU admission and/or in-hospital death was associated with being unvaccinated compared with being vaccinated. ORs were adjusted for age, geographic region, calendar time (days since January 1, 2021), and local virus circulation (percentage of SARS-CoV-2–positive results from testing within the counties surrounding the facility on the date of the encounter) and weighted for inverse propensity to be vaccinated or unvaccinated (calculated separately for each OR estimate). Generalized boosted regression trees were used to estimate the propensity to be vaccinated based on the following socio-demographic, facility, and medical factors: age, sex, race, ethnicity, Medicaid status, calendar date, geographic region, local SARS-CoV-2 circulation on the day of each medical visit, urban-rural classification of facility, hospital type, number of hospital beds, chronic respiratory condition, chronic non-respiratory condition, asthma, chronic obstructive pulmonary disease, other chronic lung disease, heart failure, ischemic heart disease, hypertension, other heart disease, stroke, other cerebrovascular disease, diabetes type 1, diabetes type 2, diabetes due to underlying conditions or other specified diabetes, other metabolic disease (excluding diabetes), clinical obesity, clinical underweight, renal disease, liver disease, blood disorder, dementia, other neurological/musculoskeletal disorder, Down syndrome, and the presence of at least one prior molecular or rapid antigen SARS-CoV-2 test record documented in the electronic medical record ≥15 days before the medical encounter date (pre-vaccination, if vaccinated). Vaccine effectiveness for prevention of COVID-19–associated ICU admission and/or in-hospital death can be estimated from the adjusted ORs presented in this table with the equation: vaccine effectiveness = (1-adjusted OR) x 100%. Adjusted ORs and VE estimates are not shown for vaccination status comparisons with confidence intervals greater than 50 percentage points around the VE estimate. Adjusted ORs and VE estimates could not be calculated for the following subgroups due to lack of model convergence: 50+ years, Moderna vaccines, 2 doses (14-149 days earlier); 50+ years, Pfizer-BioNTech vaccines, 2 doses (14-149 days earlier); all adults, heterologous vaccines, 2 doses (14-149 days earlier); all adults, heterologous vaccines, 3 doses (7-119 days earlier); 50+ years, heterologous vaccines, 2 doses (14-149 days earlier); and 50+ years, heterologous vaccines, 3 doses (7-119 days earlier). In vaccination status subgroups with <10 SARS-CoV-2–positive cases, all numbers in the row were removed because of small cell sizes. In-hospital death was defined as death in the hospital occurring ≤28 days after admission. Analyses for ICU admission and/or in-hospital death included SARS-CoV-2–positive cases with ICU admission and/or in-hospital death and all SARS-CoV-2–negative hospitalized controls. CI indicates confidence interval; ICU, intensive care unit; IQR, interquartile range; OR, odds ratio; Ref, referent group; VE, vaccine effectiveness.

**eFigure 4. Association of COVID-19–Associated Emergency Department or Urgent Care Encounters with Prior Vaccination with Two, Three, or Four mRNA Vaccine Doses, by Age Group, Among Patients Without a Prior Documented SARS-CoV-2 Infection.**

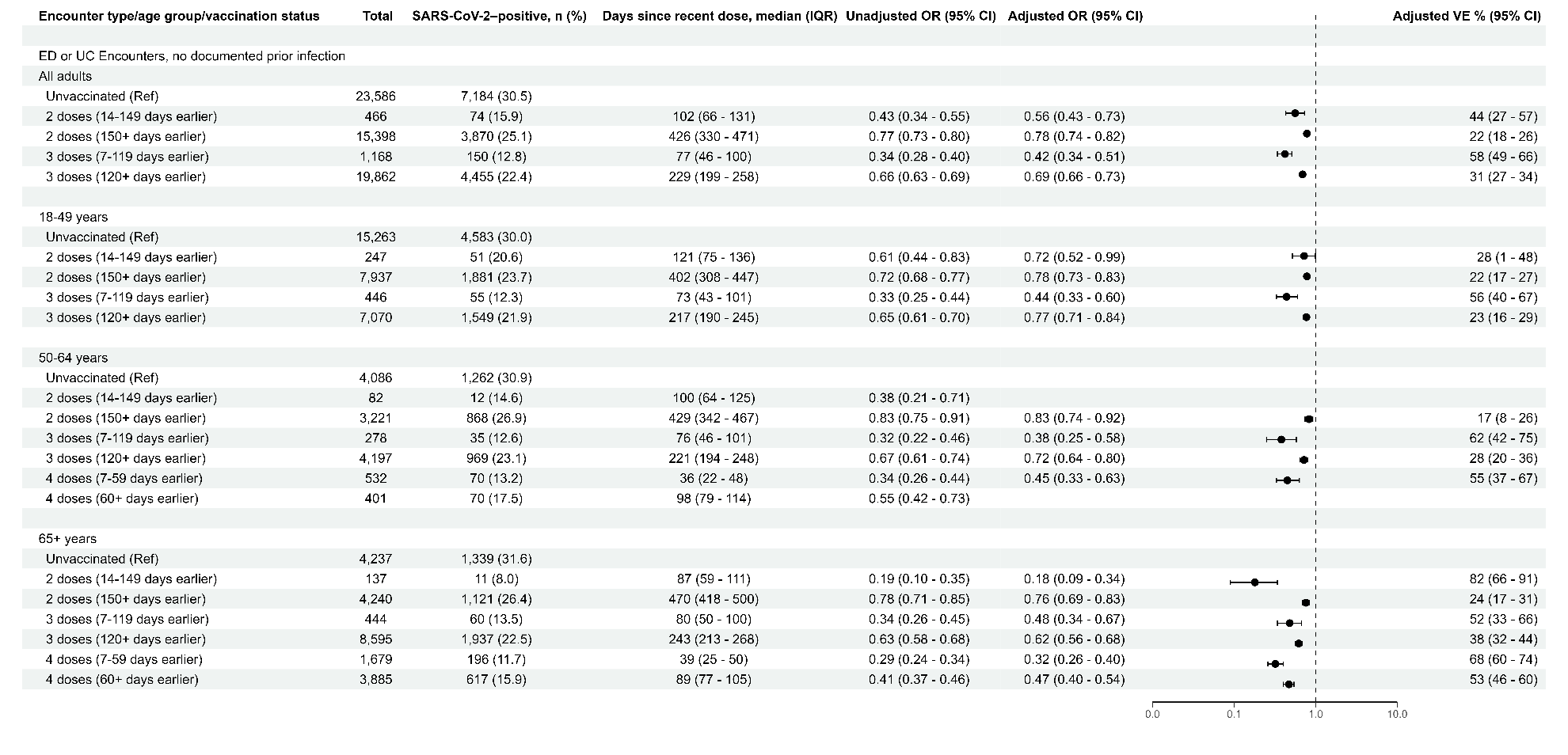

Patients included had no prior positive molecular or rapid antigen SARS-CoV-2 test result documented in the electronic medical record ≥15 days prior to the ED or UC encounter date. An adjusted OR <1.0 indicates that COVID-19–associated ED/UC encounter was associated with being unvaccinated compared with being vaccinated. ORs were adjusted for age, geographic region, calendar time (days since January 1, 2021), and local virus circulation (percentage of SARS-CoV-2–positive results from testing within the counties surrounding the facility on the date of the encounter) and weighted for inverse propensity to be vaccinated or unvaccinated (calculated separately for each OR estimate). Generalized boosted regression trees were used to estimate the propensity to be vaccinated based on the following socio-demographic, facility, and medical factors: age, sex, race, ethnicity, Medicaid status, calendar date, geographic region, local SARS-CoV-2 circulation on the day of each medical visit, urban-rural classification of facility, chronic respiratory condition, chronic non-respiratory condition, asthma, chronic obstructive pulmonary disease, other chronic lung disease, heart failure, ischemic heart disease, hypertension, other heart disease, stroke, other cerebrovascular disease, diabetes type 1, diabetes type 2, diabetes due to underlying conditions or other specified diabetes, other metabolic disease (excluding diabetes), clinical obesity, clinical underweight, renal disease, liver disease, blood disorder, dementia, other neurological/musculoskeletal disorder, Down syndrome, and the presence of at least one prior molecular or rapid antigen SARS-CoV-2 test record documented in the electronic medical record ≥15 days before the medical encounter date (pre-vaccination, if vaccinated). Vaccine effectiveness for prevention of COVID-19–associated ED/UC encounter can be estimated from the adjusted ORs presented in this table with the equation: vaccine effectiveness = (1-adjusted OR) x 100%. Adjusted ORs and VE estimates are not shown for vaccination status comparisons with confidence intervals greater than 50 percentage points around the VE estimate. CI indicates confidence interval; ED, emergency department; IQR, interquartile range; OR, odds ratio; Ref, referent group; UC, urgent care; VE, vaccine effectiveness.

**eFigure 5. Association of COVID-19–Associated Hospitalization with Prior Vaccination with Two, Three, or Four mRNA Vaccine Doses, by Age Group, Among Patients Without a Prior Documented SARS-CoV-2 Infection.**

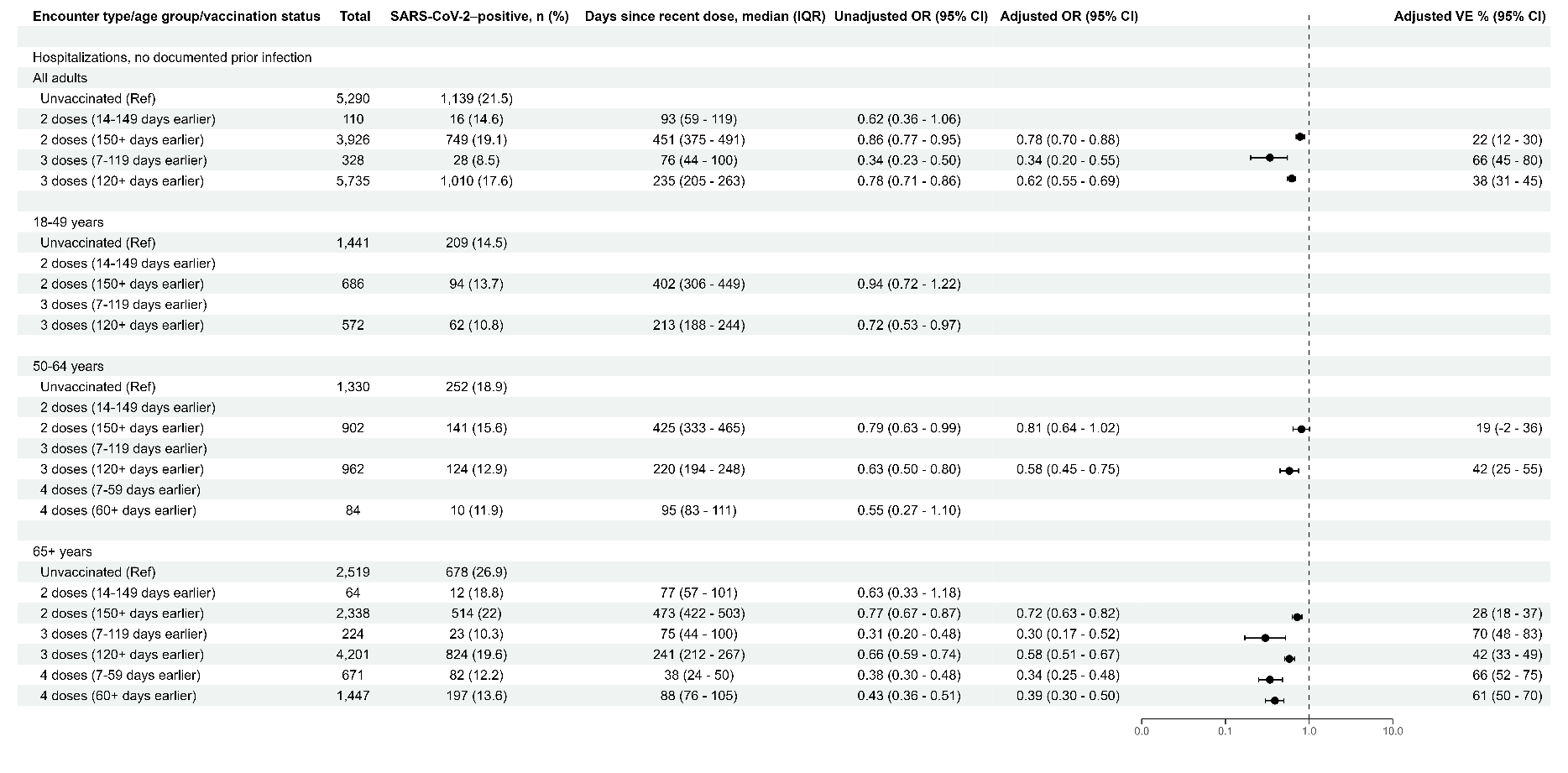

Patients included had no prior positive molecular or rapid antigen SARS-CoV-2 test result documented in the electronic medical record ≥15 days prior to the hospital admission date. An adjusted OR <1.0 indicates that COVID-19–associated hospitalization was associated with being unvaccinated compared with being vaccinated. ORs were adjusted for age, geographic region, calendar time (days since January 1, 2021), and local virus circulation (percentage of SARS-CoV-2–positive results from testing within the counties surrounding the facility on the date of the encounter) and weighted for inverse propensity to be vaccinated or unvaccinated (calculated separately for each OR estimate). Generalized boosted regression trees were used to estimate the propensity to be vaccinated based on the following socio-demographic, facility, and medical factors: age, sex, race, ethnicity, Medicaid status, calendar date, geographic region, local SARS-CoV-2 circulation on the day of each medical visit, urban-rural classification of facility, hospital type, number of hospital beds, chronic respiratory condition, chronic non-respiratory condition, asthma, chronic obstructive pulmonary disease, other chronic lung disease, heart failure, ischemic heart disease, hypertension, other heart disease, stroke, other cerebrovascular disease, diabetes type 1, diabetes type 2, diabetes due to underlying conditions or other specified diabetes, other metabolic disease (excluding diabetes), clinical obesity, clinical underweight, renal disease, liver disease, blood disorder, dementia, other neurological/musculoskeletal disorder, Down syndrome, and the presence of at least one prior molecular or rapid antigen SARS-CoV-2 test record documented in the electronic medical record ≥15 days before the medical encounter date (pre-vaccination, if vaccinated). Vaccine effectiveness for prevention of COVID-19–associated hospitalization can be estimated from the adjusted ORs presented in this table with the equation: vaccine effectiveness = (1-adjusted OR) x 100%. Adjusted ORs and VE estimates are not shown for vaccination status comparisons with confidence intervals greater than 50 percentage points around the VE estimate. Adjusted ORs and VE estimates could not be calculated for the following subgroup due to lack of model convergence: 50-64 years, 2 doses (14-149 days earlier). In vaccination status subgroups with <10 SARS-CoV-2–positive cases, all numbers in the row were removed because of small cell sizes. CI indicates confidence interval; IQR, interquartile range; OR, odds ratio; Ref, referent group; VE, vaccine effectiveness.

**eFigure 6. Association of COVID-19–Associated Intensive Care Unit Admission and/or In-Hospital Death with Prior Vaccination with Two, Three, or Four mRNA Vaccine Doses, by Age Group, Among Patients Without a Prior Documented SARS-CoV-2 Infection.**

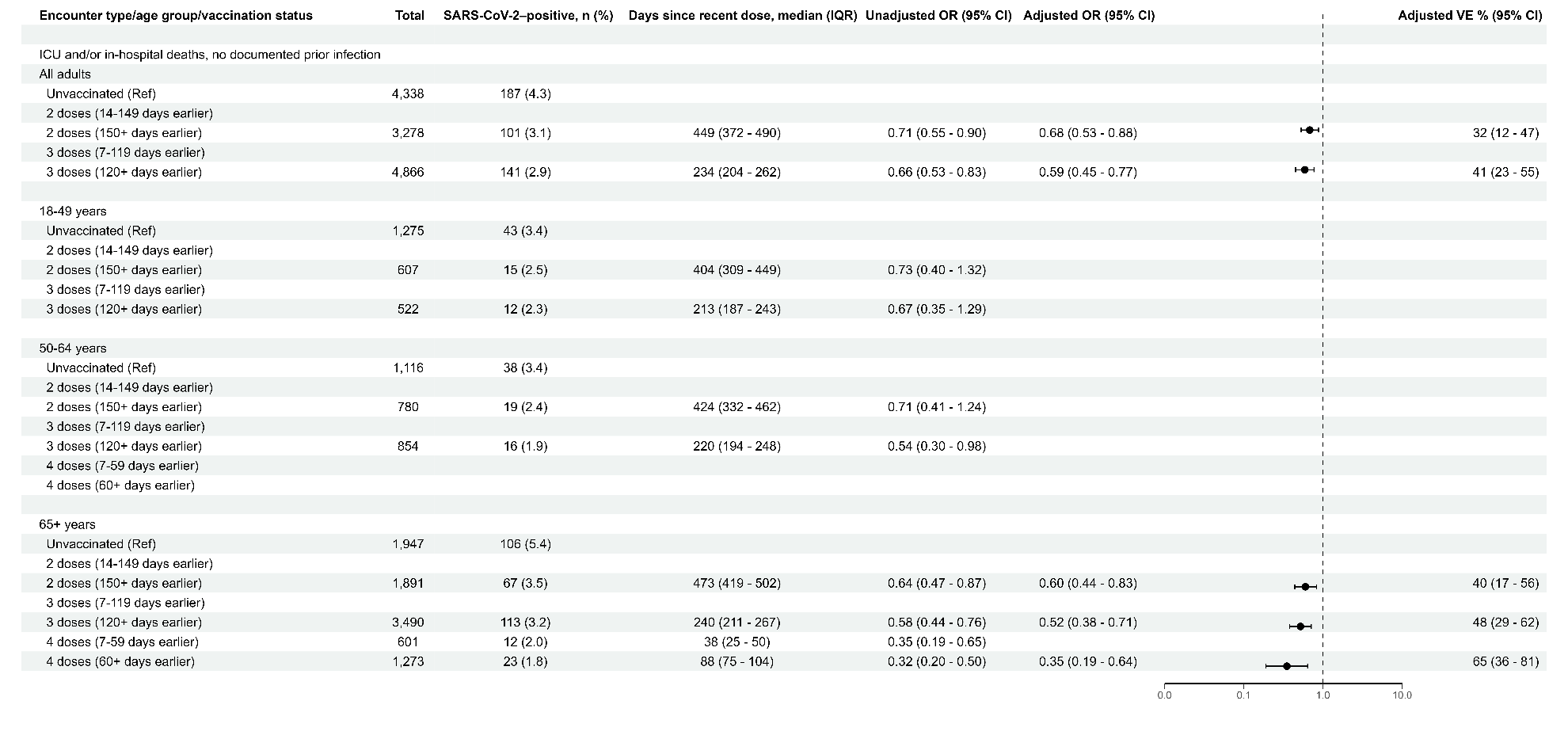

Patients included had no prior positive molecular or rapid antigen SARS-CoV-2 test result documented in the electronic medical record ≥15 days prior to the hospital admission date. An adjusted OR <1.0 indicates that COVID-19–associated ICU admission and/or in-hospital death was associated with being unvaccinated compared with being vaccinated. ORs were adjusted for age, geographic region, calendar time (days since January 1, 2021), and local virus circulation (percentage of SARS-CoV-2–positive results from testing within the counties surrounding the facility on the date of the encounter) and weighted for inverse propensity to be vaccinated or unvaccinated (calculated separately for each OR estimate). Generalized boosted regression trees were used to estimate the propensity to be vaccinated based on the following socio-demographic, facility, and medical factors: age, sex, race, ethnicity, Medicaid status, calendar date, geographic region, local SARS-CoV-2 circulation on the day of each medical visit, urban-rural classification of facility, hospital type, number of hospital beds, chronic respiratory condition, chronic non-respiratory condition, asthma, chronic obstructive pulmonary disease, other chronic lung disease, heart failure, ischemic heart disease, hypertension, other heart disease, stroke, other cerebrovascular disease, diabetes type 1, diabetes type 2, diabetes due to underlying conditions or other specified diabetes, other metabolic disease (excluding diabetes), clinical obesity, clinical underweight, renal disease, liver disease, blood disorder, dementia, other neurological/musculoskeletal disorder, Down syndrome, and the presence of at least one prior molecular or rapid antigen SARS-CoV-2 test record documented in the electronic medical record ≥15 days before the medical encounter date (pre-vaccination, if vaccinated). Vaccine effectiveness for prevention of COVID-19–associated ICU admission and/or in-hospital death can be estimated from the adjusted ORs presented in this table with the equation: vaccine effectiveness = (1-adjusted OR) x 100%. Adjusted ORs and VE estimates are not shown for vaccination status comparisons with confidence intervals greater than 50 percentage points around the VE estimate. Adjusted ORs and VE estimates could not be calculated for the following subgroups due to lack of model convergence: 18-49 years, 3 doses (7-119 days earlier); 50-64 years, 2 doses (14-149 days earlier); and 65+ years, 2 doses (14-149 days earlier). In vaccination status subgroups with <10 SARS-CoV-2–positive cases, all numbers in the row were removed because of small cell sizes. In-hospital death was defined as death in the hospital occurring ≤28 days after admission. Analyses for ICU admission and/or in-hospital death included SARS-CoV-2–positive cases with ICU admission and/or in-hospital death and all SARS-CoV-2–negative hospitalized controls. CI indicates confidence interval; ICU, intensive care unit; IQR, interquartile range; OR, odds ratio; Ref, referent group; VE, vaccine effectiveness.
